# Work Intensity and Labour Supply

**DOI:** 10.1101/2020.03.27.20041335

**Authors:** Jean Roch Donsimoni

**Author notes:** I would like to thank the members of my dissertation committee, Klaus Wälde, Florian Hett, and Philip Sauré, for their insights and comments. This work has also benefited from useful comments and discussions with Marten Hillebrand, as well as with seminar participants at Johannes Gutenberg-Universität Mainz, the University of Bath, and the 32^*nd*^ EEA annual congress in Lisbon.

## Abstract

We develop a model where individuals accumulate fatigue from work intensity when choosing hours worked. Fatigue captures intertemporal costs of labour supply and leads to a utility loss. As fatigue increases, individuals optimally choose to work fewer hours. The model also predicts that if individuals cannot easily shift consumption over time, they will work fewer hours but accumulate more fatigue when work intensity increases. Calibration to 19 European countries provides evidence for the claim that a higher share of the service sector is linked to increasing work fatigue and that public provisions of healthcare improves recovery and mental health.

**JEL codes:** E71, I12, J22

## 1 Introduction

### Motivation

There is a growing body of evidence that although average hours worked per worker are declining over time in most rich countries, various measures of stress and mental fatigue have been on the rise, where we observe increasing scores on the General Health Questionnaire which measures mental ill-health across countries and over time (see reports by OECD, 2012; 2014).^1^ We also observe cross-sectional evidence whereby countries with higher levels of sick leave from work stress generally exhibit lower average hours worked per worker (Eurostat, 2017).^2^ Standard models like Heckman (1974) and Becker (1965) cannot explain this phenomenon, as they would predict that utility costs of work should be higher when labour supply is higher. This is because in these frameworks, only contemporaneous costs of work are considered and thus working more today leads to higher current costs. Additionally, the effect does not appear to be driven by less hours worked as a direct result of sick leave. On average fewer than 4% of employees take sick leave every year according to Eurofound (2010). Meanwhile, work stress accounts only for a small fraction of total sick leave: 11.5% in the UK (Office of National Statistics, 2017) and 11.6% in Germany (Bundesanstalt für Arbeitsschutz und Arbeitsmedizin, 2016). Thus, it cannot drive the downward trend on average hours worked per worker.

### Open issue

There is little theoretical work on the effect of work-related stress and its effects on hours worked and labour supply decisions, while only some empirical evidence exists.^3^ From the perspective of work and organisational psychology, sick leave from work stress is traditionally seen as a manifestation of an individual’s increasing levels of mental fatigue as they deal with increasingly stressful work demands (see Lee and Ashforth (1996), and Wright and Cropanzano (1998)). In this case, work fatigue accumulates over time as a consequence of work effort, both at the intensive margin of work intensity and the extensive margin of hours worked. As economists, we may then consider that fatigue captures intertemporal costs of work, arising from work intensity.

Understanding the determinants of hours worked remains an important issue for the design of labour market policies, and this phenomenon of work fatigue appears to be a driver of labour supply decisions that has so far escaped the attention of economists. The traditional view that has persisted to this day is to consider only contemporaneous costs of work in the labour decision of individuals. However, given that choices of hours worked seem to have intertemporal costs and that the current thinking in labour supply cannot explain the facts mentioned above, a new framework to model labour supply decisions could resolve both issues.

### Objective

This paper offers a theoretical model explaining the negative relationship observed between work fatigue from work stress and hours worked. We will also aim to analyse the intertemporal costs of work using work fatigue as a product of stressful job demands. And finally, we will aim to give an explanation for the observed cross-sectional stylised fact about the link between sick leave rates and hours worked at country level.

### Framework

The individual in the baseline model maximises lifetime utility, where consumption equals labour income.^4^ Over time, work fatigue increases with hours worked and with work intensity. Work fatigue also depreciates exogenously, capturing the individual’s ability to recover or the efficiency of healthcare provisions in dealing with work fatigue. Thus, the consumer faces a trade-off in their labour decision: when choosing hours worked, they can increase their current consumption via labour income, but this will also increase fatigue over time. In turn, fatigue negatively affects the utility function of the consumer as a stock variable capturing effects of previous as well as current labour supply decisions. This modelling choice replaces the traditional ‘disutility of labour’ by a ‘disutility of work fatigue’.

In modelling fatigue accumulation, we take into account that work intensity (i.e. stressful work demands from the individual’s job) is the factor causing hours worked to increase fatigue (Demerouti *et al.*, 2001). We consider that work intensity can be measured by the number and the complexity of the tasks that a worker has to perform in their job, e.g. writing a new paper, teaching a new course, or applying for a large multinational research grant. The recovery of the individual will take place at a constant rate and be proportional to the current stock of fatigue.^5^ Thus, we rule out individual agency in the recuperation from fatigue, e.g. by investment into recuperation technologies or strategies. Instead, we focus exclusively on recovery from exogenous sources. The last element of the model is the production sector with a representative firm with constant total factor productivity using labour services as their only input into production.

### Findings

There are two main contributions of this paper. First, we show that as fatigue increases, the policy function for labour supply will be decreasing. We therefore show that there exists a negative relationship, at the individual level, between hours spent working and fatigue from work intensity. Extending the model to consider wealth and human capital does not affect this relationship. Relatedly, we also find that if consumption is more inelastic over time, long-run hours worked will decrease while long-run fatigue will increase when work intensity goes up.

Second, we calibrate the model to match a cross-section of 19 European countries, showing a negative relationship between sick leave rates from work stress and aggregate full-time hours worked. We let the sensitivity of hours worked to stressful demands and the depreciation rate of fatigue adjust to hit targets for hours worked and sick leave rates from work stress, respectively. The distribution of calibrated parameters for the former can be explained by an increasing share of the service sector in the economy, offering evidence for the claim from psychology that countries with higher shares of services should experience higher work fatigue (Schaufeli *et al.*, 2008)^6^, here proxied by sick leave rates from work stress. Meanwhile, the distribution of parameters for the depreciation of fatigue can partly be explained by increasing shares of government health expenditure in primary care services out of total health expenditure. This supports the view that public provisions of healthcare help reduce the negative effects of work stress and aid in the recovery mental health issues (Layard, 2016).

### Related literature

This paper fits within the tradition of two important strands of economic research: the literature on health capital and the demand for health, started by Grossman (1972), and the theoretical literature on dynamic models of labour supply, pioneered by Heckman (1974). The closest paper to our work in the literature on the demand for health services come from French (2005), who studies the effects of social security benefits and health status on retirement behaviour and labour supply in a structural model. In his paper, the author finds that calibrating the model against a panel data set of US households, he can explain both life-cycle patterns of labour supply and labour force participation, and is able to identify key determinants to retirement behaviour at older ages. However, although the model can simulate labour supply and labour force participation patterns accurately up to age 60, the later years are often underestimated. Labour supply of unhealthy individuals late in life is substantially larger in the model than in the data, while labour force participation of unhealthy individuals is significantly and substantially larger in the model than in the data throughout the life-cycle. Our model has some key differences to the model by French (2005). First, as in Grossman (1972), French (2005) uses health status as a positive good in the utility function, with no relation to labour supply. In our framework, we instead propose the notion that it is exactly labour supply that drives current and future (mental) health status. Second, the costs of work in French (2005) are contemporaneous costs of work, as is customary in the labour literature, while we consider these costs to be intertemporal and capture not only the costs of work in the current period but also in previous periods. Furthermore, our framework could help to understand labour supply decisions toward the end of the life-cycle for unhealthy individuals, as these individuals could potentially exhibit large levels of work fatigue from stressful demands accumulated over their career, leading them to work much fewer hours than predicted by standard models.

This paper is also part of the literature on dynamic labour supply. This literature finds its roots in the papers of Heckman (1974) and Heckman and MaCurdy (1980), who first formalised the now standard Neoclassical view whereby individuals maximise lifetime utility, with a contemporaneous trade-off between consumption and leisure. Since then various extensions have been proposed, from the inclusion of human capital accumulation (Heckman, 1976; Blinder and Weiss, 1976) to the consideration of individual characteristics such as marriage decision and fertility (Heckman and Willis, 1977; Weiss and Gronau, 1981). More recently, efforts have been made to link micro and macro theories of labour supply. Most recently, Erosa *et al.* (2016) have proposed a life-cycle model with heterogeneous agents in a small-open economy. Individuals in the model are heterogeneous in their labour productivity, based on their education level, and can transition out of the labour market with a given probability every period, for which they are compensated by unemployment insurance. The authors closely match life-cycle patterns of labour force participation and unemployment spells. However, their model overestimates hours worked later in life, near retirement, especially for highly educated workers. Thus, in a departure from this literature, and the work of Erosa *et al.* (2016), our framework will seek to capture the phenomenon that work choices have intertemporal costs for the utility and that work intensity, as measured by the amount of stressful job demands, is a powerful force behind it. This will lead us to capture drops in labour supply in the long-run, which may serve to explain why most life-cycle models of labour supply often overestimate hours worked late in life.^7^

More specifically, and pertaining to the question at hand about the link between hours of work and mental health in the form of work fatigue, we note the contributions of Cygan-Rehm and Wunder (2018) and Banerjee *et al.* (2017). Cygan-Rehm and Wunder (2018) find that when hours worked increase, both objective and subjective health measures decrease. In particular, mental health is more affected than physical health, landing ever more support to the idea that we must understand the link between labour supply decisions and mental health. Banerjee *et al.* (2017) find further evidence in the same direction. They find that declining mental health adversely affects labour force participation and increases sick leave and absenteeism. Lastly, we also acknowledge the contribution of Lang *et al.* (2019), who find that increased import competition worsens both physical and mental health measures over time, with the latter being a more persistent effect. They use the accession of China to the WTO to study how increased competition from Chinese manufacturers affected the well-being of US manufacturing workers. Their proposed channel is that of negatively affected labour market outcomes, namely employment, wages, and job security. The dimension of work intensity is not explored explicitly. However, the mechanism of increased competition harming domestic firms implies a reduction in work intensity as demand for goods domestically produced decreases. In our paper, we choose to focus on this channel of work intensity in a closed economy.^8^

### Structure

Section 2 introduces the baseline model and discusses optimal behaviour. The first part of this section will focus on a general form of the model, before using specific functional forms to derive more precise implications. Section 3 discusses labour-fatigue dynamics for the individual, by exploring both steady-states and transitional dynamics and provides a simple calibration to support the findings of the baseline model. Section 4 offers an application of the model to a stylised fact, offering evidence for the observed variation across countries in hours worked and sick leave rates from work stress. Section 5 discusses possible limitations of the model in terms of its application, and provides two extensions to the baseline model. Finally, Section 6 concludes.

## 2 The Baseline Model

In this section we describe the baseline model of labour supply with fatigue as a measure of the utility costs of work. Extensions are presented later on in Section 5, addressing potential concerns over the assumptions of the baseline model. Allowing for savings as well as for human capital investments does not change the main result of the paper.

### 2.1 The representative firm

Time is continuous and indexed by *t* ∈ [0, ∞). The production sector in the economy is characterised by a representative firm producing the final good *Y* (*t*) ∈ ℝ_+_. The firm uses (aggregate) labour inputs *L* (*t*) ∈ ℝ_+_ as their sole factor of production. The production function takes the following linear form, *Y* (*t*) = *AL* (*t*), where *A* ∈ ℝ_+_ is constant labour productivity. Thus, output in this economy grows at the same rate as labour. The firm maximises current profits taking the wage as given. The usual result then emerges that the first-order condition of the firm implies that factor rewards equal their marginal productivity. This in turn implies that, on aggregate, the wage rate will be constant and equal to labour productivity *A*.^9^

### 2.2 Consumer problem and optimal behaviour

We describe the problem of a single household within the economy. Later, we explore the dynamics of the model for a given individual *I* ∈ {1, 2, …, *N*}, where we assume a constant population size *N* > 0. We will detail later the dimension along which individuals differ.

#### 2.2.1 Work fatigue from work intensity

This section describes the accumulation of work fatigue, captured by *g*_*i*_ (*t*) ∈ ℝ_+_, as a function of hours worked and the current stock of fatigue at time *t*,

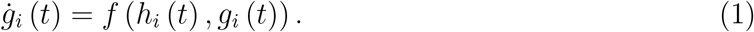

Fatigue is defined on the set of non-negative reals, the reasoning behind this being that ‘negative’ fatigue has no clear interpretation. Individuals can either be psychologically exhausted or not, and if so, the question then is to what degree they are exhausted. In this setup, the presence of stressful job demands makes the function f (·) more responsive to changes in labour supply 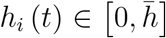, taken to represent the total time endowment the individual can choose from at any point in time *t* for an individual *i*, with 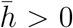. On the other hand, a higher depreciation rate of fatigue would make *f* (·) more responsive to changes in fatigue *g*_*i*_ (*t*). This depreciation of the stock of fatigue can be interpreted either as the worker’s ability to cope with and recover from fatigue at the individual level or as the quality of the institutions granting healthcare services for mental health issues (e.g. stress from work) at the aggregate level. We make the following assumptions about changes in the stock of fatigue.

##### Assumption 1.

*The function f* : 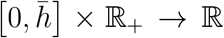 *in C*^2^ *both of its arguments, and is strictly increasing convex in h*_*i*_

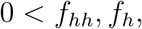

*and strictly decreasing in g*_*i*_

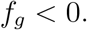

*Furthermore, f is additively separable in h*_*i*_ *and g*_*i*_, *meaning that the cross-partial derivatives are equal to zero*

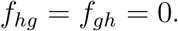

Firstly, hours worked have a strictly non-negative marginal effect on the growth rate of fatigue. This allows us to rule out situations where working somehow reduces the negative effect of work fatigue, which could include situations such as individuals alleviating their stress or even recuperating through working. This would be inconsistent with the hypothesis that fatigue increases with exposure to stressful demands on the job as work intensity increases. Furthermore, hours worked are assumed to have an increasingly negative impact on the growth rate of fatigue by convexity. This implies that, ceteris paribus, the more hours an individual works, the faster their fatigue will accumulate. For example working one more hour from 12h/day to 13h/day will have a greater effect on fatigue accumulation than working one more hour from 2h/day to 3h/day.

Secondly, the change in fatigue is assumed to be strictly decreasing in the stock of fatigue. This means that fatigue depreciates over time, independently from investments in labour supply. It captures the ability of individuals (or in the aggregate, the efficiency of the healthcare system) to deal with fatigue. This allows us to make the following remark.

##### Corollary 1.

*Given Assumption 1, there exists a point* 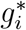 *such that* 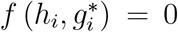, *and if* 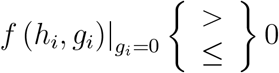 0, *then we have* 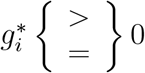.

This simplifies the analysis by making the steady-state level 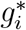 stable and unique. Having a unique steady-state level of fatigue 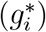 may appear at first to be a strong assumption, however it would be more surprising if the individual had multiple optimal levels of fatigue. Also, we assume that the marginal effect of hours worked on fatigue accumulation does not depend on fatigue, while the marginal effect of fatigue on its own change does not depend on hours worked. That is, we separate the recovery process from the effects of stressful job demands. Allowing for non-zero cross partial derivatives would imply that the ability of the individual to recover from fatigue would somehow be dependent on their work environment and the amount of hours worked. It would also imply that the effect of work demands would depend on the level of fatigue of the individual as well as on their recovery capabilities. While plausible, we choose to abstract from these considerations.

#### 2.2.2 Preferences and utility loss from work fatigue

Consumers derive utility from consuming goods at time *t*, denoted *c*_*i*_ (*t*) ∈ ℝ_+_, while experiencing disutility from work fatigue at time *t, g*_*i*_ (*t*). Work fatigue is positively correlated with higher levels of anxiety, sleep disturbance and cognitive impairments (Peterson *et al.*, 2008), but also with lower life satisfaction and depression (Hakanen and Schaufeli, 2012). These negative effects on individual well-being are captured below in (2) and serve to illustrate the utility loss from work fatigue. The instantaneous utility function is of the form

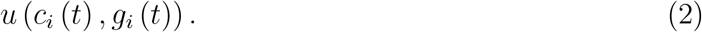

We make the following assumptions on the behaviour of the utility function.

##### Assumption 2.

*The function* 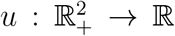 *is C*^2^ *in both of its arguments, and is strictly increasing concave in c*_*i*_

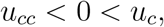

*and decreasing concave in g*_*i*_

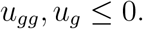

*Concavity of u in g*_*i*_ *captures the fact that as work fatigue increases, its effects on individual well-being become increasingly more severe.*

In the model presented here, work fatigue accumulates over time as a reaction to exposure to stressful tasks at work, and as it accumulates, its strain on individual utility increases at an increasing rate. Thus we model only the direct effect of fatigue on the individual, and abstract from indirect effects such as cognitive impairments, which may also affect productivity via a separate channel.

Intertemporal preferences are given by the following objective function giving lifetime utility as the discounted sum of instantaneous utility as given in (2), where *ρ* ∈ ℝ_+_\{0} is the time preference rate,

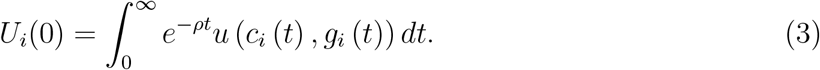

We assume that the individual cannot save or borrow,^10^ and consumption is therefore limited to equal labour income,^11^ with wage *w* ∈ ℝ_+_\{0},

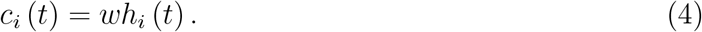

The individual chooses labour supply *h*_*i*_ (*t*), which will pin consumption *c*_*i*_ (*t*) by the budget constraint.

#### 2.2.3 Optimal rule for labour supply

The individual maximises discounted lifetime utility from (3) subject to (1) and (4), yielding the optimal rule for labour supply (see app. A.1 for steps)

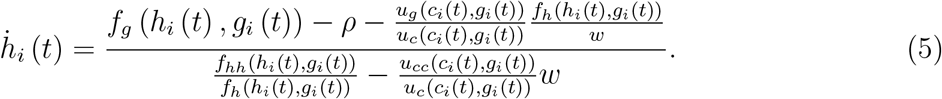

We can see that the only force driving the growth rate of hours worked upward is a compensatory motive determined by the ratio of marginal utilities, since 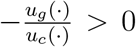. This term shows the trade-off in the utility of increasing hours worked between the marginal loss from fatigue and the marginal gain from consumption. This term is multiplied by the ratio of the marginal effect of hours worked on the growth rate of fatigue, *f*_*h*_ (·), and the wage *w*, which captures the trade-off in the constraints of increasing labour supply. We summarise this result in Lemma 1.

##### Lemma 1

(Compensatory motive). *Individuals will increase their hours worked faster over time when marginal disutility from fatigue increases in order to compensate for the marginal utility loss, relative to the marginal utility from consumption.*

This shows that individuals internalise the costs of fatigue and will increase their hours worked faster, in order to compensate for the increasing relative marginal disutility from work fatigue. The marginal effect of *u*_*g*_ (·) on the growth rate of hours worked is measured by the inverse of the marginal utility of consumption, as well as the relative cost of hours worked on the accumulation of fatigue over the wage, functioning as a real measure of an increase in hours worked on fatigue. We can further see that the wage rate will have a negative effect in levels both in the numerator and in the denominator of (5) as it increases. Finally, we can see from (5) that *ρ* and *f*_*g*_ (·) will both have a strictly non-positive effect on the growth rate of hours worked. As the individual values the present more, i.e. as *ρ* increases, they will choose a lower labour supply path, while an increase in the marginal effect of fatigue on its on growth rate, i.e. a greater rate of depreciation of fatigue, will also put the individual on a lower labour supply path. This system of equations completely characterises the model in equilibrium.

#### 2.2.4 Modelling preferences and fatigue explicitly

We will now take the system of equations above as described by (1), (4), and (5) and consider a specific functional form for (2). The utility function is now given by

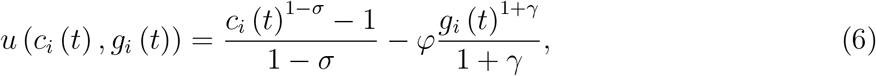

where *σ* ∈ ℝ_+_\{0} is the inverse of the intertemporal elasticity of substitution [IES], *φ* ∈ ℝ_+_ is a weight parameter for the relative weight of the utility loss from fatigue, and *γ* ∈ ℝ_+_ is the elasticity of fatigue. We also specify a functional form for the accumulation of work fatigue from (1),

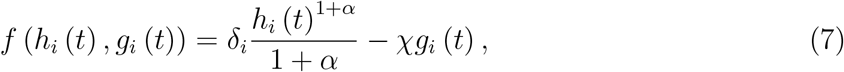

where the first term on the right-hand side of (7) captures the effect of stressful job demands *δ*_*i*_ ∈ ℝ_+_\{0}, and the second term models the recovery process, with *χ* ∈ ℝ_+_. In later sections, when we consider changes in work intensity *δ*_*i*_. Furthermore, we introduce ex-ante heterogeneity by having individuals draw a level of work intensity *δ*_*i*_ from a known distribution. We can understand it as a recent graduate applying for jobs and being allocated to a job with intensity *δ*_*i*_ with probability *p*_*i*_. This could happen because after applying to a large number of positions, the recent graduate only receives a few offers but has no ability to discern the level of work intensity in each job. After the draw, the worker then faces a deterministic problem. Thus, because of the heterogeneity in *δ*_*i*_, the paths of hours worked and fatigue will differ across individuals.

On the other hand, *α* ∈ ℝ_+_ measures how elastic hours worked are in the accumulation of fatigue, and thus how prone a particular type of job may be to accumulating work fatigue. Differences between individuals in *α* may arise because individuals working in different industries may perform jobs that on average are more prone to accumulating work fatigue. Their job could involve dealing with customers, students, or patients, which would make any given work hour more emotionally draining than working with machinery.

We further assume that, at the individual level, the worker has a natural physical and mental ability to recover from, and cope with, their own fatigue. The aggregate analogue would be the efficiency of the healthcare system in providing services to deal with and recover from work-related fatigue from stressful job demands. Thus, the individual can be either more resilient, i.e. high *χ*, or more susceptible, i.e. low *χ*, to fatigue, as a result of different institutional backgrounds for healthcare services.

The choice of recuperation, or coping, strategies are not modelled here and cannot be endogenously chosen by the individual in the current framework. The assumption is that the worker can only affect how much labour they supply while letting fatigue depreciate naturally through their own coping capabilities.

Using (6) and (7) in the system described by (1) and (5), we can use (4) to replace consumption in (5) and rearrange to obtain the following system of equations,

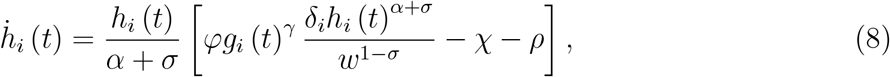

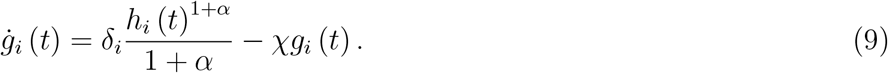

These two equations now describe the reduced form of the system explicitly. This allows us to study both the dynamics and the long-run behaviour of individuals in the next section.

### 2.3 Aggregation

After workers draw a *δ*_*i*_ from an arbitrary distribution, with *i* = 1, 2, …, *N*, they keep it for the rest of their working life. The rest of the model remains otherwise deterministic. This in turn implies that aggregate labour supply will be given by 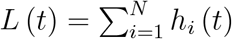. Other aggregate variables are defined analogously. By applying a standard law of large numbers and letting *N* grow sufficiently large, this means that labour supply per capita will be equal to average labour supply

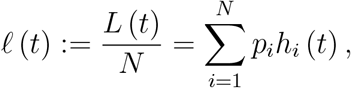

where *p*_*i*_ is the probability of having drawn *δ*_*i*_ at the start of the planning horizon which, using a law of large numbers, corresponds to population shares. This implies that the growth rate of average labour supply is equal to the weighted mean of the growth rates of individual labour supply,

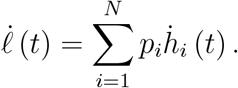

Multiplying by *N* gives the growth rate of aggregate labour supply. The growth rates of aggregate consumption and aggregate fatigue are defined similarly. In the next section, we focus on a single individual’s behaviour to gain insight into the dynamics of the model, before returning to the macroeconomy in the extensions.

## 3 Are Long Work Hours Optimal?

### 3.1 Working less for more fatigue in the long-run

#### Steady-state

Our system of two equations in two variables can easily be solved to obtain equilibrium steady-state values for labour supply and fatigue. These steady-state values for hours worked and fatigue are summarised in Lemma 2, and allow us to understand the channels affecting the long-run behaviour of individuals.

##### Lemma 2

(Steady-state). *Let ξ* ≡ *α* + *σ* + *γ* (1 + *α*), *then we can write the steady-state level of hours worked in closed-form solution as*

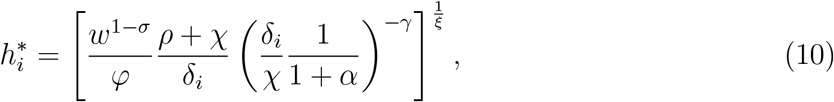

*and the corresponding steady-state level of fatigue as*

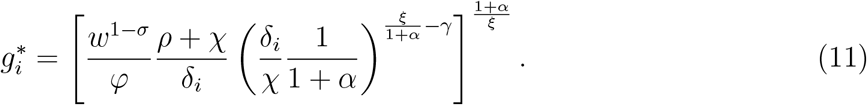

We can see from expressions (10) and (11) that the determinants of hours worked and fatigue in the steady-state will be similar. Specifically, we can see that the ratio of the workload *δ*_*i*_ to the recuperation rate *χ*, weighed by the gross elasticity of fatigue 1 + *α* in hours worked will have different effects depending on whether one looks at hours worked or fatigue. In the case of the former, it will have a negative effect owing to the elasticity of utility in fatigue *γ*. While the latter will be pushed upwards due to the sign of its exponent being positive, 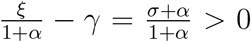. This already hints at a negative relationship between hours worked and work fatigue in the long-run. On the other hand, the wage rate *w*, time preference rate *ρ*, and weight of the utility loss from fatigue *φ* will all affect steady-state values of labour and fatigue similarly. Comparative-statics analysis of the steady-state values for labour supply and work fatigue gives rise to the following proposition.

#### Lemma 3

(Comparative statics).

i. *The long-run value of labour supply* 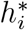 *increases with: (a) a higher depreciation rate of fatigue χ, (b) a higher time preference rate ρ, (c) a lower weight of the utility loss from fatigue ϕ, (d) lower levels of stressful job demands δ*_*i*_, *and (e) if σ* < 1, *a higher wage rate w.*
ii. *The long-run value of work fatigue* 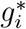 *increases with: (a) a higher depreciation rate of fatigue χ, (b) a higher time preference rate ρ, (c) a lower weight of the utility loss from fatigue ϕ, (d) a lower elasticity of fatigue in the utility function γ, and (e) if σ* < 1, *a higher wage rate w and lower job demands δ*_*i*_.

This is intuitive to understand. (i.a) If fatigue depreciates faster, individuals recover more quickly, which means they can afford higher fatigue levels in the long-run, pushing allowing workers to work longer hours. (i.b) In this model, impatience translates into a desire to consume more today, which is accomplished here by working longer hours at the cost of higher fatigue. (i.c) If the weight of the utility loss from fatigue drops, the individual suffers less from the adverse effects of labour supply in the long-run and thus chooses to work more. (i.d) Lower job demands imply a slower accumulation of work fatigue, meaning once again that the individual can afford to work longer hours. (i.e) When the substitution effect dominates the income effect, workers will want to take advantage of their higher labour income induced by higher wages and choose to work more hours.

The effects on the long-run value of fatigue are also intuitive. (ii.a) Because fatigue depreciates faster, the individual chooses to work more hours (as they can recover more quickly), sustaining a higher level of fatigue in the long-run, from which they know they can recover more rapidly. (ii.b) Similarly, because individuals are impatient, they incur more fatigue as a direct consequence of working more. (ii.c) As in the case of higher depreciation, lower weight of the utility loss implies higher long-run fatigue, however in this case it is because the individual simply does not experience a sufficient negative response to discourage higher fatigue accumulation. (ii.d) When the utility loss from fatigue is less responsive to changes in fatigue, the individual reacts less strongly, causing them to accumulate more fatigue because their preferences do not react fast enough to increasing fatigue. (ii.e) As the substitution effect dominates the income effect, the individual works more and accrues more fatigue. The secondary effect, regarding lower job demands, is driven by the opportunity cost of work changing with the IES. When job demands decrease, it makes the gains from higher hours worked larger, which the individual chooses to exploit, leading them to accumulate more work fatigue in the long-run. This last point offers a surprising result.

##### Proposition 1.

*Steady-state fatigue* 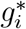 *increases in work intensity δ*_*i*_ *iff the IES is smaller than one, i.e.*

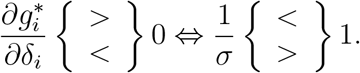

*Proof.* Taking the derivative of (11) w.r.t. work intensity yields the result immediately

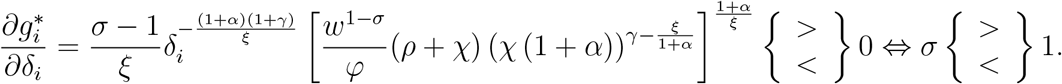

▪

The intuition behind this result is an unexpected insight from the model. When a worker finds it difficult to shift consumption over time, steady-state fatigue will increase when work intensity increases, despite steady-hours worked decreasing.^12^ The only mechanism in the model consistent with this result is for the individual to select a steeper labour path, accumulating fatigue rapidly by working high hours. Thus, when work becomes more demanding, we have the result that despite working less in the long-run, this type of worker will face greater utility costs and higher fatigue.

#### Transitional dynamics

In order to analyse the transitional dynamics of the model, we must look at the two-dimensional system in *h*_*i*_ and *g*_*i*_. We proceed by taking the derivative of hours worked with respect to work fatigue, cancelling out the change over time in both dynamic rules. Thus, we consider fatigue as the exogenous variable using the system in (8)-(9).

##### Definition 1

(Optimal path). *The optimal behaviour of individuals in the labour-fatigue space is determined entirely by the following expression*,

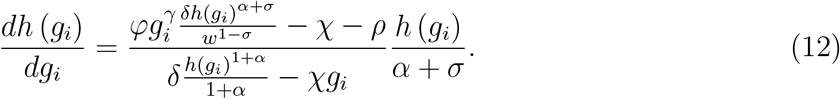

*Furthermore, along the equilibrium path to the steady-state, the transversality condition [TVC] holds (see app. A.3)*,

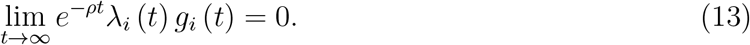

In the definition above, *λ*_*i*_ (*t*) is the associated shadow price of fatigue. This condition ensures that fatigue cannot grow too fast compared to its marginal value fixed by the shadow price *λ*_*i*_. This condition is equivalent to saying that the individual cannot gain by deviating from the optimal path implied by equation (13) and never returning to it.^13^ Thus the TVC, combined with the expression in (12), gives us the optimal path for hours worked and fatigue leading to the maximal level of lifetime utility the individual can achieve.

#### Proposition 2.

*The optimal path to the steady-state will be characterised by* 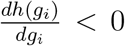. *In other words:*

i. *Individuals will work more hours and reduce their labour supply as fatigue increases if* 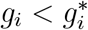,
ii. *while individuals will work fewer hours and increase their labour supply as they recover if* 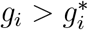.

*Proof.* See app. A.2.

▪

Proposition 2 summarises the main message of Figure 1 and the key result of this paper. We can see that the optimal behaviour of agents in the model will lead to a negative relationship between labour supply and work fatigue, supporting the earlier stylised fact from country cross-sections.

**Figure 1:**
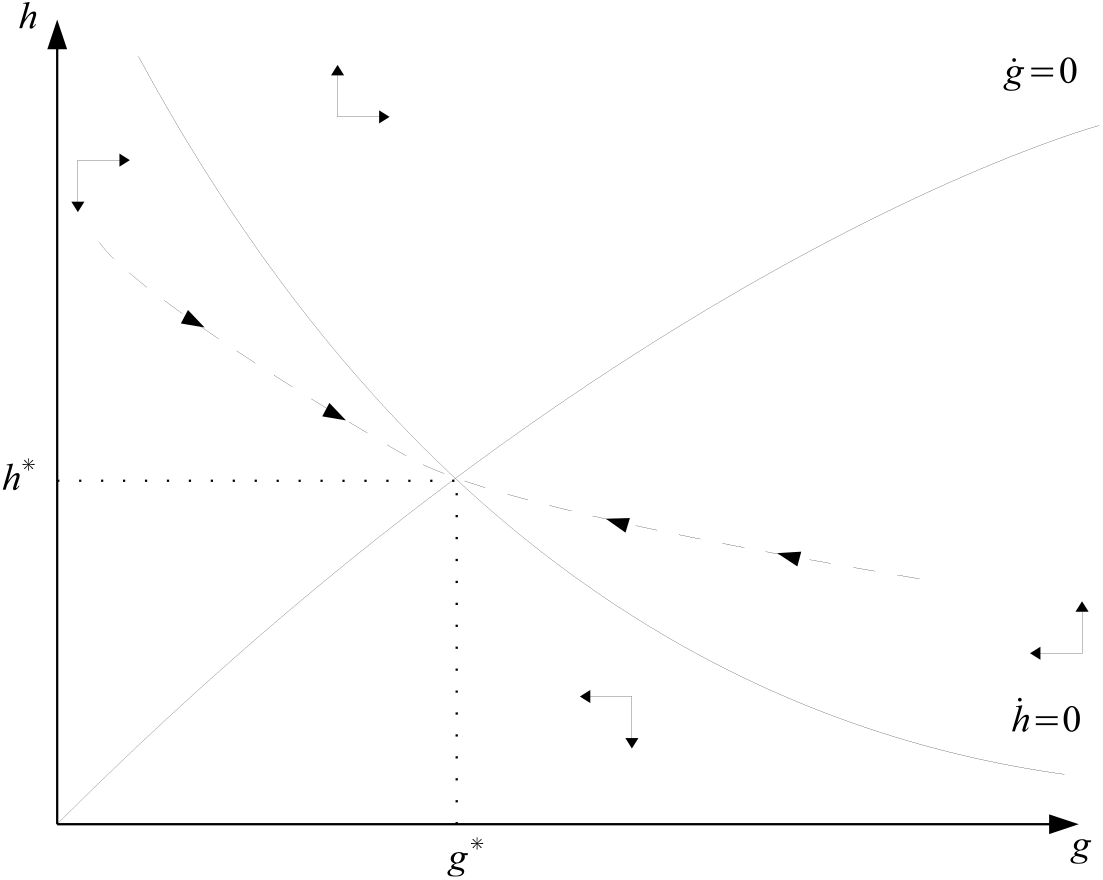
Dynamics of labour supply and fatigue along the stable saddle-path

When fatigue is low, individuals will work more and progressively reduce their labour supply as work fatigue accumulates. This behaviour is driven by individuals taking advantage of their low level of fatigue to work more and thus be able to consume more and maximise utility. However, as fatigue begins to settle in and take its toll on the individual’s well-being, they will reduce their hours worked. This occurs as the utility loss from fatigue becomes large compared to the utility gains from consumption.

On the other hand, if fatigue is high, individuals will work fewer hours, in order to allow themselves to recover from work fatigue. They will gradually increase their labour supply as their mental health improves, until they can reach their long-run value for hours worked and fatigue, characterised by the steady-states 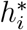 and 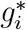. Deviations from the optimal path, though prevented by the TVC, would lead individuals to over accumulate fatigue if they chose a higher path of labour supply. Should they choose a sub-optimal amount of hours worked, then they would never become fatigued, and eventually they would reduce hours worked to zero. The question now remains to know how long individuals take to reach an arbitrarily close neighbourhood of their steady-state, and if this behaviour can be confirmed by numerical methods for reasonable parameter values and targets.

### 3.2 Overtime at a cost: a quantitative illustration

We proceed with an illustrative calibration exercise in order to ascertain whether the dynamics predicted by our baseline model are consistent with empirical observations, given known parameter values from the literature.

#### 3.2.1 A simple calibration

##### Defining the targets

We pause here to address the fact that the targets and parameter values are for averages of aggregate values at country level, while using the consumer problem only. We now focus our attention on a representative consumer, and thus now abstract from concerns over heterogeneity in work demands, causing heterogeneous levels of labour supply and work fatigue. In this case, the optimal behaviour of individuals will be identical to the aggregate behaviour of all workers in the economy and we drop the subscript *i* from this point on. This is due to the definition of aggregate labour supply as summing the hours worked of all workers in the economy, 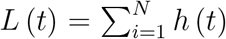. Since all individuals are identical, there is no stochastic process, and ex-ante heterogeneity has been removed, we can rewrite this expression as *L* (*t*) = *Nh* (*t*), assuming all individuals have the same initial conditions.^14^ This implies that the growth rate of individual and aggregate variables are identical. We now return to discussing the parameter set and targets below.

In order to obtain a meaningful solution for hours worked and fatigue, parameters are chosen so as to obtain steady-state values that are consistent with real world observations for hours worked and work fatigue. To this end, we use Germany as an example, targeting average weekly hours worked per worker and mean effort scores for measures of 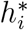 and 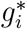 respectively. We note here that we use average annual hours worked per worker and then scale it down to weekly hours worked.

The Effort-Reward Imbalance [ERI] inventory (Siegrist *et al.*, 2004) consists of two main scales used to assess individuals level of accumulated fatigue from work effort and the level of rewards they receive (financial and non-financial). Respondents answer various questions about their work, divided along the lines of an Effort scale and a Reward Scale. We choose here to focus on the Effort scale as it is the most directly related measure of work fatigue from job demands. The ERI effort scale is made up of six items scored between 1 and 5, giving a total score between 6 and 30, where the higher the score the more work effort the individual spends.

The targets are a long-run value of hours worked of 39.8h/week, corresponding to average hours worked for Germany in 2015 (OECD, 2018), and an ERI effort score of 13.8, corresponding to the general population average in Germany, also for 2015 (Hinz *et al.*, 2016). We calibrate the fatigue propensity of a job, *α*, to hit the long-run value of hours worked, and the depreciation rate of fatigue, *χ*, to hit the long-run value of work fatigue.

#### Calibrating

We use values for the IES, the time preference rate, the wage rate, and work intensity taken directly from the literature. We assume values of *σ* = 0.74, taken from Andersen *et al.* (2008) and *ρ* = 4.507%, taken from Gourinchas and Parker (2002). The values chosen imply that the substitution effect will dominate the income effect, as *σ* < 1, which will lead individuals to choose paths consistent with higher preferences for current gains from hours worked. While the consumer discounts the future at a rate of 4.6% annually.^15^ We also set the annual wage rate to the average annual wage in Germany in 2015, and set *w* = €39, 218 (OECD, 2018a). The last parameter to be exogenously set is the level of stressful job demands *δ*. We set it using the measure of work intensity from Avgoustaki and Frankort (2019), which they obtained using the 2010 and 2015 waves of the European Working Conditions Survey. They define a composite measure of work intensity using various item responses from the survey with values ranging from 0 to 6, with 0 implying the individual never faces any intense job demands and 6 meaning they always face this type of demands. We use the average value of work intensity, using their composite indicator, found in the cross-European sample over the two waves, and set *δ* = 2.69. The last two parameters, *γ* and *φ* are free parameters and simply adjusted to obtain meaningful solutions.

We obtain the following results from our calibration exercise, with values of *α* = × 1.95589 10^−4^ and *χ* = 0.0406907, while hitting our targets exactly. These values imply that the type of work consistent with the targets is one that has low sensitivity to stressful demands, and that the degree of provisions of healthcare services toward recuperating from work fatigue (or alternatively, the ability of individuals to cope with work fatigue) is relatively low.

#### 3.2.2 Policy function and time paths

The numerical analysis begins by showing the saddle-path to equilibrium from Figure 1 quantitatively, using the parameter set from Table 1. We therefore look at Figure 2 to show the numerical counterpart to the phase diagram in the previous section. The optimal path to the steady-state declines monotonically in fatigue. It leads to a value of labour supply of 39.8h/week, corresponding to average weekly hours worked, and a value for fatigue of 13.8, corresponding to average scores for individuals in full-time employment on the ERI effort scale.

**Table 1:** Exogenous and calibrated parameter values with numerical targets for Germany in 2015

| Parameter | Value | Definition | Source |
| --- | --- | --- | --- |
| *Exogenous* |  |  |  |
| $w$ | 39,218 | average annual wage in (EUR) | OECD (2018a) |
| $\rho$ | 0.04507 | time preference rate | Gourinchas and Parker (2002) |
| $\sigma$ | 0.74 | inverse of IES | Andersen *et al.* (2008) |
| $\delta$ | 2.69 | work intensity | Avgoustaki and Frankort (2019) |
| $\gamma$ | 1 | elasticity of utility loss from $g$ | Free |
| $\varphi$ | 0.11519 | weight of utility loss from $g$ | Free |
| *Calibrated* |  |  |  |
| $\chi$ | 0.0406907 | depreciation rate of fatigue | calibrated |
| $\alpha$ | $1.95589 \times 10^{-4}$ | fatigue propensity of job | calibrated |
| *Targets* |  |  |  |
| $h^*$ | 39.8 | Average FT hours worked/week | OECD (2018b) |
| $g^*$ | 13.8 | Average ERI effort score | Hinz *et al.* (2016) |

**Figure 2:**
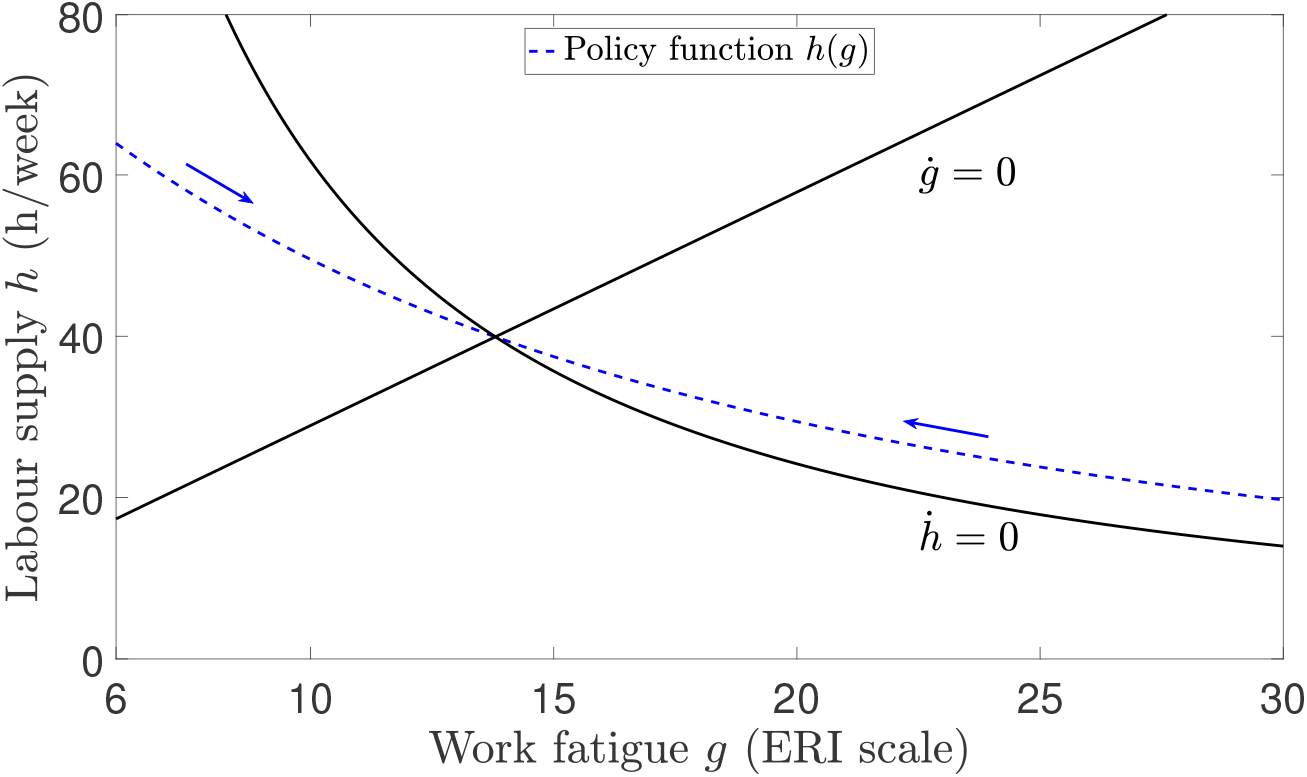
Hours worked per week decline as work fatigue increases

Therefore we can see that the numerical analysis confirms the analytical solution derived previously and shows a substantial shift in hours worked depending on the level of work fatigue. Indeed, the phase diagram shows that when fatigue is lowest, individuals will work over 60h per week.^16^ If on the other hand the individual begins to the right of the steady-state, where their work fatigue level is high and hours worked are low, then they will progressively adjust their work hours upward while they recover and their fatigue reduces. We see that they would work around 20h per week, corresponding to part-time hours worked, with work fatigue levels of 30, or the maximum possible score. Of course, this quantitative phase diagram does not offer any insight about the time paths of labour supply or fatigue. For this, we turn to the next figure.

Figure 3 plots the system characterised by (8)-(9) over time, defined by years on the job, or tenure. The time steps taken are in years, while the left-hand scale shows average hours worked per week and the right-hand scale shows fatigue, as measured by the ERI effort scale. Individuals at the beginning of the planning horizon will work close to 65 hours per week when work fatigue is at its lowest point (i.e. 6 on the ERI effort scale). However, when facing a constant wage, hours worked will decline and within 10 years drop by more than 15 hours per week before slowing down and converging to the steady state value of 39.8 hours per week over the remainder of the time horizon. Thus, we can see that while the decline of labour supply as fatigue increases is smooth in Figure 2, the bulk of the transition downward in hours worked occurs in the first decade of the working life of individuals.

**Figure 3:**
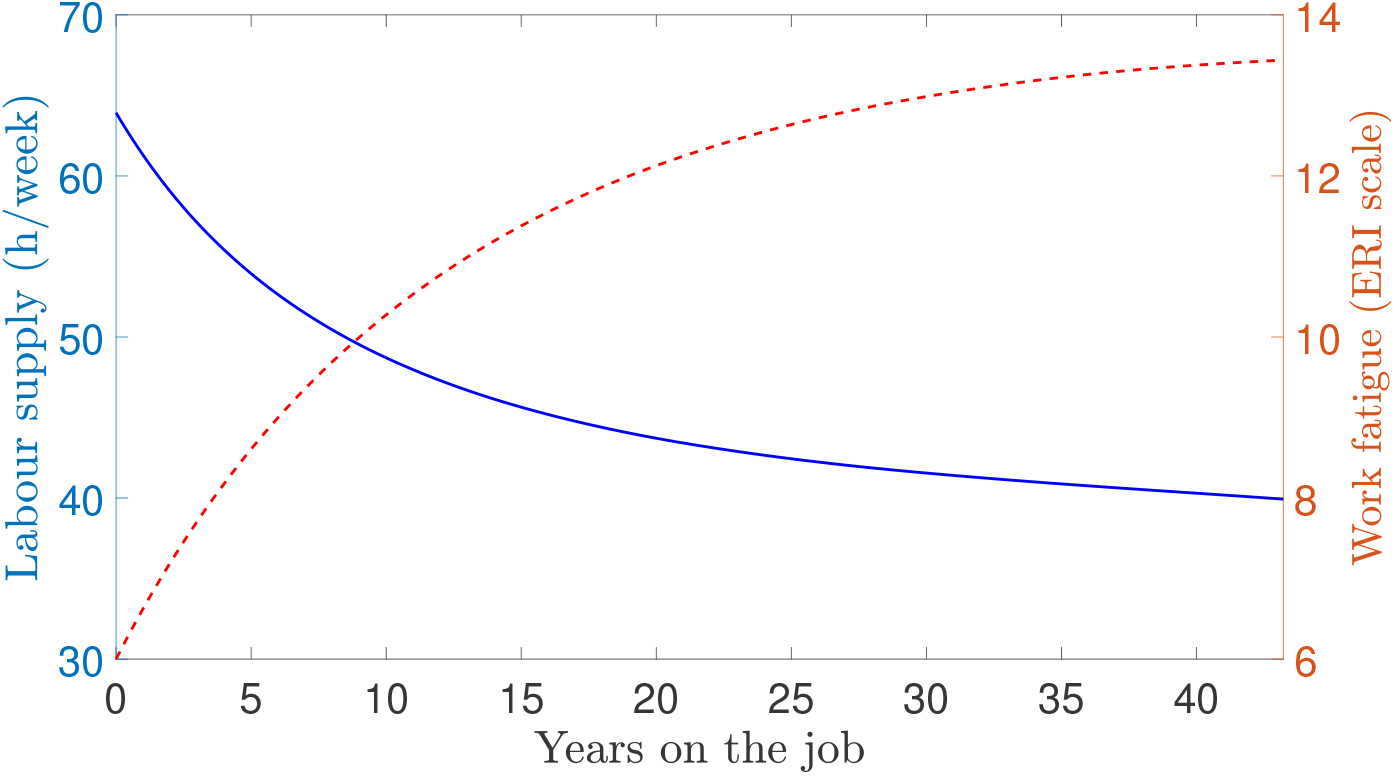
Hours and fatigue profiles over time converge to their steady-state values

Meanwhile, work fatigue will accumulate and converge to its steady state value within 45 years. Thus, as individuals start their working lives with no work fatigue, they can afford to work longer hours and maximise their labour income, which corresponds exactly to consumption. The progressive accumulation of work fatigue on the individual will lead them to reduce their hours worked over time to reduce the utility loss, converging towards their long-run values.

### 3.3 Long-run effects of institutions

We can look at the effect that “institutional” parameters of the model will have on the long-run behaviour of individuals by charting their effects on the steady-state values of labour supply and work fatigue. Figures 4(a)-4(d) show how the steady-state point represented by (*g*^*^, *h*^*^) moves inside of the labour-fatigue space when the institutional parameters of the model change. The figures show that the effects of the institutional background will vary significantly, depending on which aspect is being considered, which may help to explain cross-country variations in the long run.

**Figure 4:**
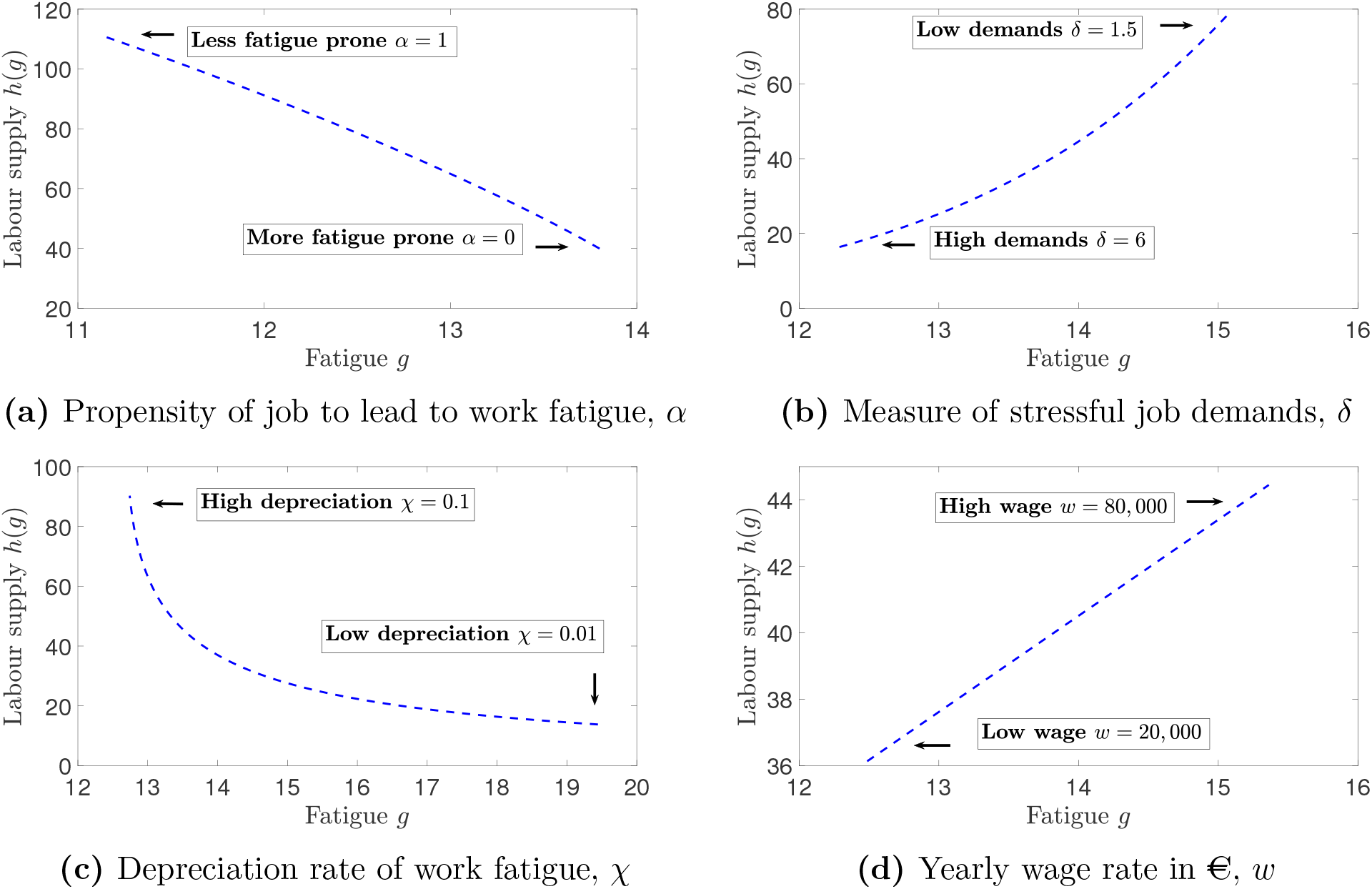
Differentiated effects of institutional background on steady-state values *h*^*^ and *g*^*^

We can see that movements from the northwest to the southeast of the phase diagram occur when either the type of work goes from less to more prone to fatigue (Figure 4(a)) or when the depreciation rate of work fatigue decreases (Figure 4(c)). This would suggest that in the long-run, countries with different levels of healthcare provisions toward mental health issues would be spread along a declining slope in the graph plotting hours worked against work fatigue. Thus, countries with more efficient healthcare services may not experience lower work fatigue but display large differences in hours worked, while on the other hand countries with less efficient healthcare services may exhibit large variations in long-run work fatigue but little differences in long-run hours worked.

Jobs that are more prone to accumulating work fatigue such as in the service sector will be scattered along a negatively slopping linear trajectory in the labour-fatigue space. Figure 4(a) shows that the type of work and how susceptible it is to work fatigue from stressful work demands will lead to large cross-country variations in both labour supply and fatigue.

We can also see that the effects of stressful job demands and the wage rate go in opposite trajectories, moving in the labour-fatigue space from southwest to northeast. We see from Figure 4(d) that even a four-fold increase in the wage rate from €20, 000 to €80, 000 per annum leads to little variation in labour supply. We can note here that this effect occurs because we have a positive wage elasticity of labour supply in our current framework. The wage elasticity of labour supply in the steady-state *h*^*^, which is exactly the term being plotted here, is obtained from equation (10) by taking logs and then differentiating with respect to *w*. This elasticity is given by

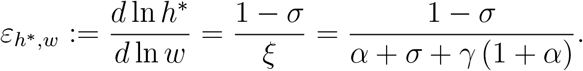

This term is positive here, since we assume a value of *σ* that is less than one.^17^ In this case, individuals are more willing to shift hours worked forward in time to enjoy potentially higher labour income in the future. Thus, in the long-run, as the wage increases they will converge toward higher levels of labour supply.

As to the effect of stressful job demands, we can see from Figure 4(b) that more stressful demands will, other things equal, reduce both the long-run level of hours worked and work fatigue. In this type of economies, consumers would seek to reduce the effects of these stressful demands by working fewer hours and, by the compensatory motive discussed in Lemma 1, will also need to reduce their fatigue in order to minimise the impact on their utility. Thus, country differences along a positive slope in the labour-fatigue space can be explained, partly, by differences in the wage rate and the measures of stressful job demands.

## 4 Application

### 4.1 The link between work stress and hours worked

We now understand the dynamics of labour supply and the intertemporal utility costs associated with stressful job demands. Thus, we are in a position to offer some insights regarding the earlier stylised fact concerning the negative relationship between average aggregate hours worked full-time and sick leave rates from work stress. We can see clearly the apparent negative correlation between sick leave from work stress and labour supply in a cross-section of European countries in 2013 (see Figure 5). We note here that the same pattern emerges when looking at data from 2007.

**Figure 5:**
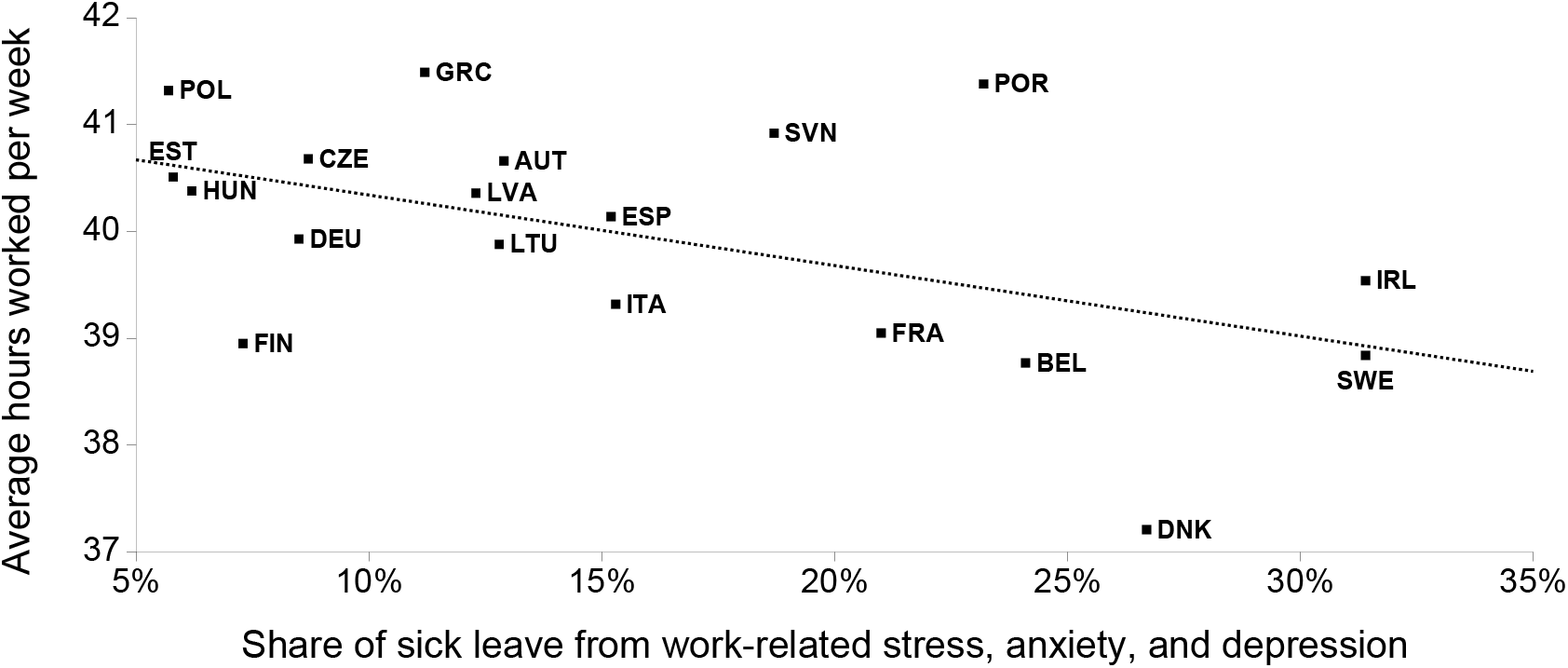
Hours worked for full-time employees fall with sick leave from work stress (Eurostat, 2017). The trend line shows the fitted values of a (simple) linear regression, the *p*-values are reported in parentheses. With slope/coefficient = −6.6^**^ (*p* = 0.025), and intercept/constant = 41^***^ (*p* = 0.000). The coefficient of determination is given by R^2^ = 0.26, and the sample size is *N* = 19.

Proposition 2 implies that we can understand this pattern as being the result of the negative association between hours worked and work fatigue. The result is understood as the optimal choice of individuals to work long hours before work fatigue settles in and progressively reduce their labour supply as they incur the costs of those past work decisions. The figure below shows the share of total sick leave caused by work-related stress, depression and anxiety in 2013 against average weekly hours worked in 2013. These measures are both aggregates, and the latter is obtained by scaling average annual hours worked, which corresponds to total hours worked in each country divided by the total number of workers.

Upon first looking at Figure 5, the casual observer may consider that the pattern is driven exactly because individual workers in Sweden, Denmark, and Ireland take more sick leave, thus reducing average hours worked in a given year. However, when looking more closely at the data, we can observe that sick leave, for all causes (including work stress) only affects less than 9% of all workers in the EU (Eurofound, 2010). Sick leave from work stress accounts for at most a third of total sick leave. Thus, even in Sweden and Ireland, this means that at most 3% of the working population is affected by sick leave from work stress. In Germany or Finland, the figure would drop to less than 1%. Thus, it would appear unlikely that these figures are driven by an argument that more sick leave from work stress induces lower average hours worked.

Finally, even assuming that all countries are identical and that no other channel is driving the data, Figure 5 does imply, when put in conjunction with the phase diagram of the model (both qualitative and quantitative), that all countries find themselves along the saddle-path to the steady-state. This would imply that each country differs only in their initial conditions and that all are converging to the same steady-state. Alternatively, it could be explained by differences in the fundamentals of their economies while all countries are in or near their steady-state. This would then mean that any change witnessed between the 2007 and 2013 cross-sections are the result of changes to those institutions rather than the result of standard transitional dynamics.

### 4.2 Does improving healthcare matter?

As mentioned above, there can be many reasons behind the country differences seen in Figure 5. These range from healthcare system design, to mental health provisions, cultural differences, or types of work. The list is not exhaustive. Our next move, therefore, is to map the parameters of the model described in the previous section to the institutional background described here, bearing in mind that only certain aspects can be captured.

In equation (7) we can note three parameters of interest: stressful job demands *δ*, the elasticity of fatigue accumulation in hours worked *α*, and the depreciation rate of fatigue *χ*. The first parameter, the measure of how stressful or fatiguing a job is will vary across countries, reflects variations in sectoral compositions, where each sector may have different levels of stressful demands. The second parameter, capturing how quickly the growth rate of fatigue (i.e. the intertemporal costs of work) increases with hours worked, captures the idea that for a given level of stressful demands *δ* faced by a (representative) worker, the impact on the increase in fatigue will be higher if hours worked are higher. This can capture differences in the type of work faced by workers, such as differences across sectors, while *δ* would capture the absolute level of work demands in a given job. The greater *α* is, the more responsive the growth rate of fatigue is to hours worked. The third, and last, parameter is the depreciation rate of fatigue. We already discussed the potential aggregate implications of this parameter earlier, where *δ* was described as encompassing efficiency of the healthcare system and the level of healthcare provisions for mental health issues relating to work stress. Thus, a more liberal healthcare system with benefits for workers suffering from job stress and work fatigue would be reflected in the model by a higher depreciation rate of fatigue *χ*. Finally, an important source of variation in hours worked across countries may come from productivity and wages. Long-run changes caused by these institutional parameters are already shown in Figures 4(a)-4(d).

Given our analysis from Section 3.2, we can focus on the effects of the propensity of the job to work stress, *α*, and the depreciation rate of fatigue, *χ*, on long-run labour supply and work fatigue. Figures 6(a) and 6(b) provide a more disaggregated picture of the effects of *α* and *χ*. Each figure plots the direct effect of an increase in each parameter on *h*^*^ and *g*^*^ separately. Here, we can see that when jobs are more prone to accumulating stress, i.e. when *α* increases, we will observe longer hours worked together with lower levels of fatigue. The effect of *α* on both steady-states is monotonic and appears to be almost linear. On the other hand, when healthcare provisions toward work stress are more effective, captured by an increase in the depreciation rate of fatigue *χ*, we observe a similar pattern for labour supply, and of similar magnitude. However, the effect on long-run fatigue is more pronounced and non-linear. Figure 6(b) shows that most of the effect of improving healthcare aimed at work stress relief will take place when moving from very little efficacy, a low *χ* near 0 to a moderate level of efficacy, represented by a value of *χ* of less than 0.03.^18^ Between these values, long-run work fatigue will drop by nearly 25% (from 20 to 15), while the remainder of the effect as *χ* continues to increase shifts work fatigue downward by 15% (from 15 to ca. 12.8).

**Figure 6:**
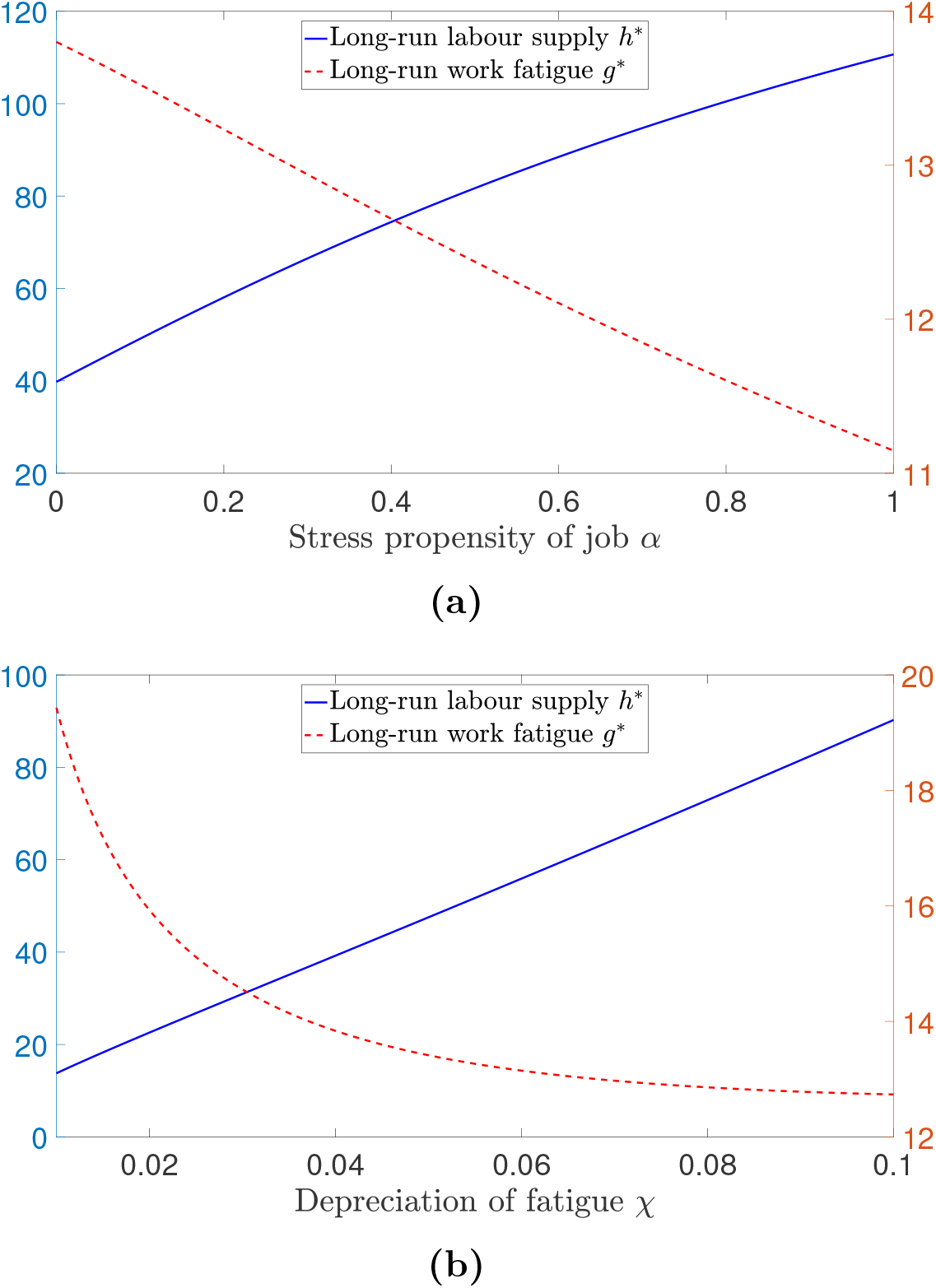
The type of job and the effectiveness of healthcare provisions in dealing with work stress have substantial, and similar, effects on labour supply but affect work fatigue differently

Given these comparative statics, it appears that affecting the healthcare system, here captured by changes in *χ*, would indeed have the largest effect on steady-state fatigue. In other words, improving the healthcare system matters.

### 4.3 What the model can tell us about the data

Using a similar approach as in the quantitative phase diagram of Figure 2, we now choose to calibrate the model to match hours worked and sick leave rates for each European country in Figure 5 for 2013. In other words, now that we have a quantitative understanding of the dynamics of labour and fatigue over time, we apply the model to a cross-section of countries to test the model against the stylised fact mentioned in the introduction.

We calibrate the model by choosing job sensitivity *α* and the depreciation rate of fatigue *χ*, in order to hit targets for hours worked and sick leave rates for each country. We compute sick leave rates by a simple affine transformation of the long-run value of fatigue *g*^*^. As argued earlier, work fatigue and sick leave rates from work stress (as a % of total sick leave) are strongly correlated phenomena.^19^

For each country, we target long-run hours worked *h*^*^ and a linear transformation of long-run work fatigue *g*^*^, approximating sick leave rates. Each country is further distinguished by wages. We use the average annual wage in Euros, in purchasing power parity in 2013 (OECD, 2018a). The rest of the parameters are as in Section 3.2.

We proceed by calibrating parameter pairs *α* and *χ* to hit both of our targets. As before, *α* is used to calibrate hours worked while *χ* is used to calibrate sick leave rates. The data for the targets is taken directly from Eurostat (2017) for each country. Figure 7 shows the comparison between the data (blue circle) and the model (red cross) values for hours worked and sick leave rates. The calibration leads us to hit most of our targets exactly, with the only exception being Finland. For this country, the model over predicts sick leave rates, given the parameter set.

**Figure 7:**
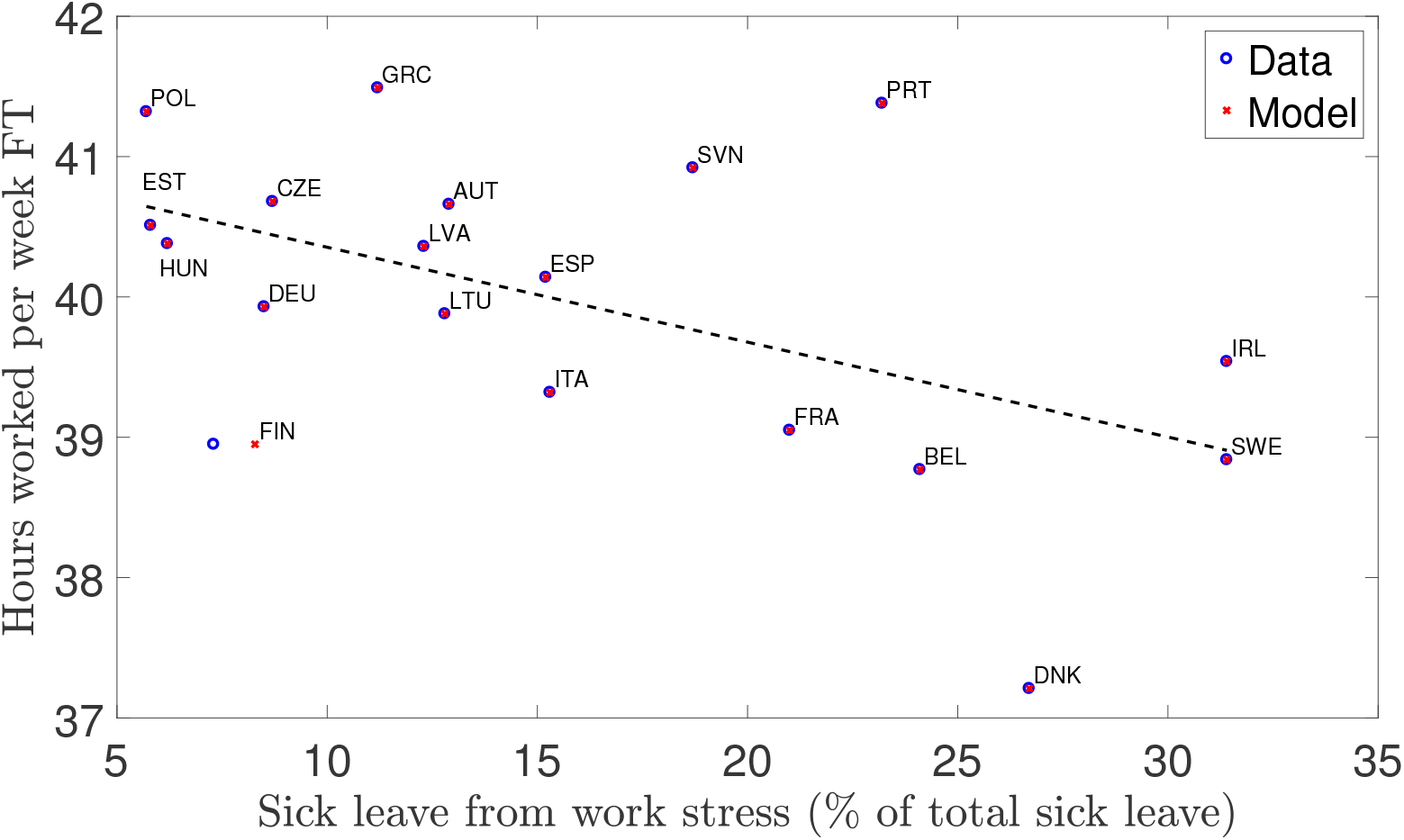
Predicted sick leave rates for Finland are higher than in the data while all other countries match exactly. The trend line shows the fitted values for the data, as in the previous scatter plot.

Firstly, since we can see from Figures 6(a) and 6(b) that both *α* and *χ* have similar effects on hours worked and fatigue, we need to check that the calibrated values are not obtained because of correlated values across both parameters. We would not want our calibration to give us both high values of *χ* and *α* whenever hours worked are higher and sick leave rates are lower. We would also like to avoid the opposite argument and have values of *α* and *χ* balance each other out in opposite directions in order to adjust to the targets. Looking at the correlation between the two vectors of calibrated values, we see that it takes a value of −0.06, which supports the idea that the calibration of one parameter does not systematically affect the calibration of the other.

Turning to the calibrated values of *α*, we see that these will range from near 0 to near 0.8 see Figure 8. This parameter is intended to capture the degree of sensitivity of the type of job to work stress. Thus, countries with higher values of *α* should be characterised by jobs that are more sensitive to work intensity in the form of stressful demands. According to organisational psychology, it has been observed that the service sector has a strong link with higher exposure to stressful demands (see e.g. Roelen *et al.* (2015) for an overview of the literature). Thus, we would expect that countries with larger service sectors should also exhibit higher values of *α*. We can plot the calibrated values of *α* from our model against the service sector share for each country in value-added, as a % of GDP (OECD, 2019b).

**Figure 8:**
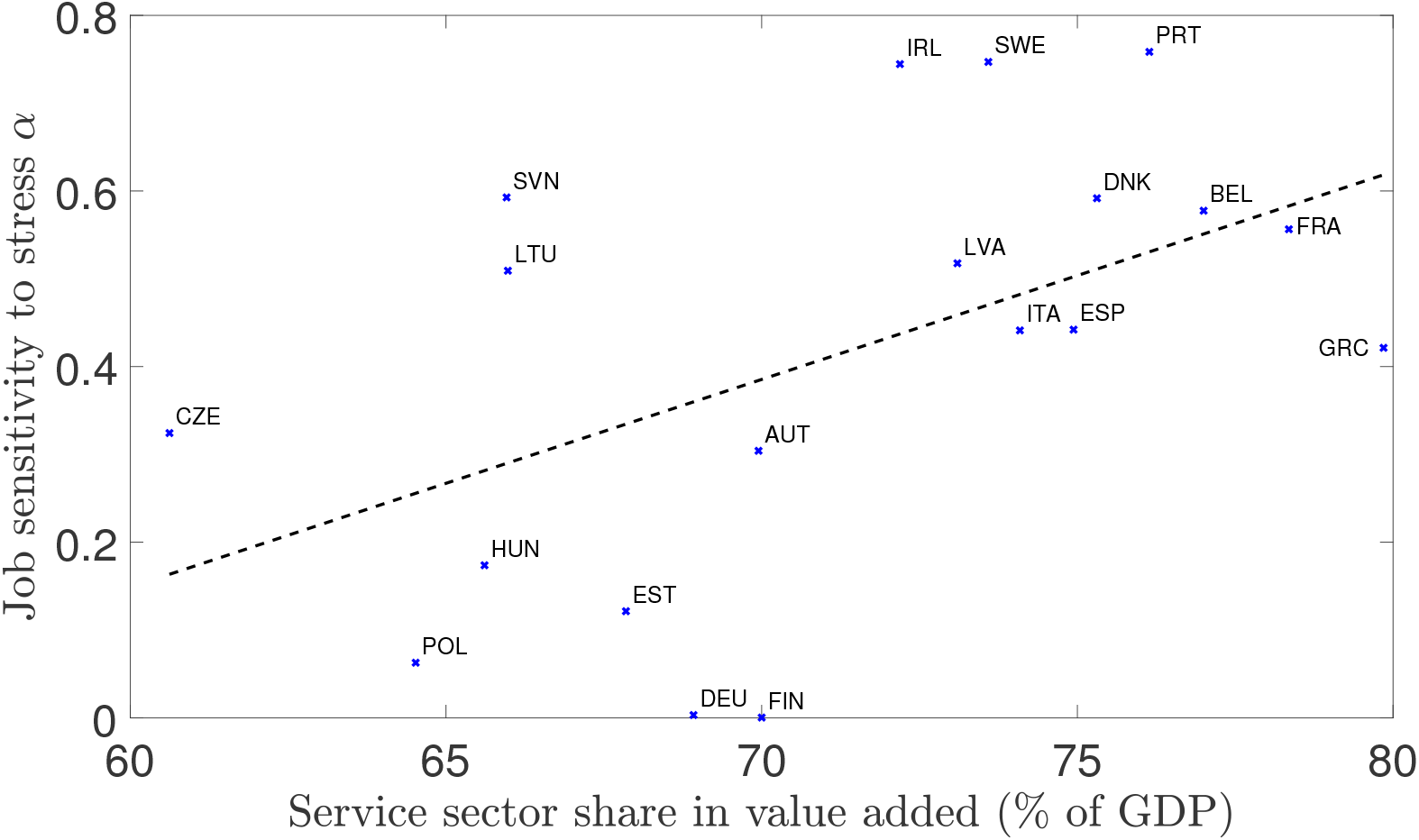
Calibrated values of *α* can be rationalised by the share of the service sector in GDP in value-added. The trend line shows the fitted values of a (simple) linear regression, the *p*-values are reported in parentheses. With slope/coefficient = 2.37^**^ (*p* = 0.028), and intercept/constant = −1.27^*^(*p* = 0.089). The coefficient of determination is given by R^2^ = 0.25, and the sample size is *N* = 19.

Figure 8 shows that the size of the service sector does appear to explain the cross-country variation in calibrated values of *α*. Germany and Finland appear to be outliers, lying near the lower bound on the domain of *α*. However, the remaining countries all seem to point into the same direction: larger service sectors correlates with higher stress sensitivity of work *α*. We can see that the fitted values are significant (see *p*-values in brackets in Figure 8) while R^2^ for the linear regression is around 0.25, despite a small sample size of 19 countries.

Next, we turn to our second calibrated parameter, the depreciation rate of work fatigue *χ*. This parameter captures the extent of the recovery from work fatigue that individuals experience at each point in time. Therefore, if a country displays a larger value of *χ*, it should be more efficient at coping with and recovering from work fatigue. Also, since *χ* is exogenous, this efficiency must be outside of the decision-making process of the individual. To capture this effect, we look at expenditures on primary care services (curative and rehabilitative services in particular, i.e. coping and recovery) from compulsory and government funded sources for each country as a share of total health expenditure (OECD, 2019a). This measure satisfies both the requirement that it should be exogenous to the decision problem of the individual and that it covers coping and recovery experiences.

Figure 9 displays the distribution of calibrated *χ* values against publicly funded primary care (as a % of total health expenditure). We can once more observe a positive relationship between our calibrated parameter set and the target measure from the data. However, we can see that the fitted values, although with a value of R^2^ of 0.12, do not appear to be significant as neither the constant nor the slope of the trend have *p*-values lower than 0.1.^20^

**Figure 9:**
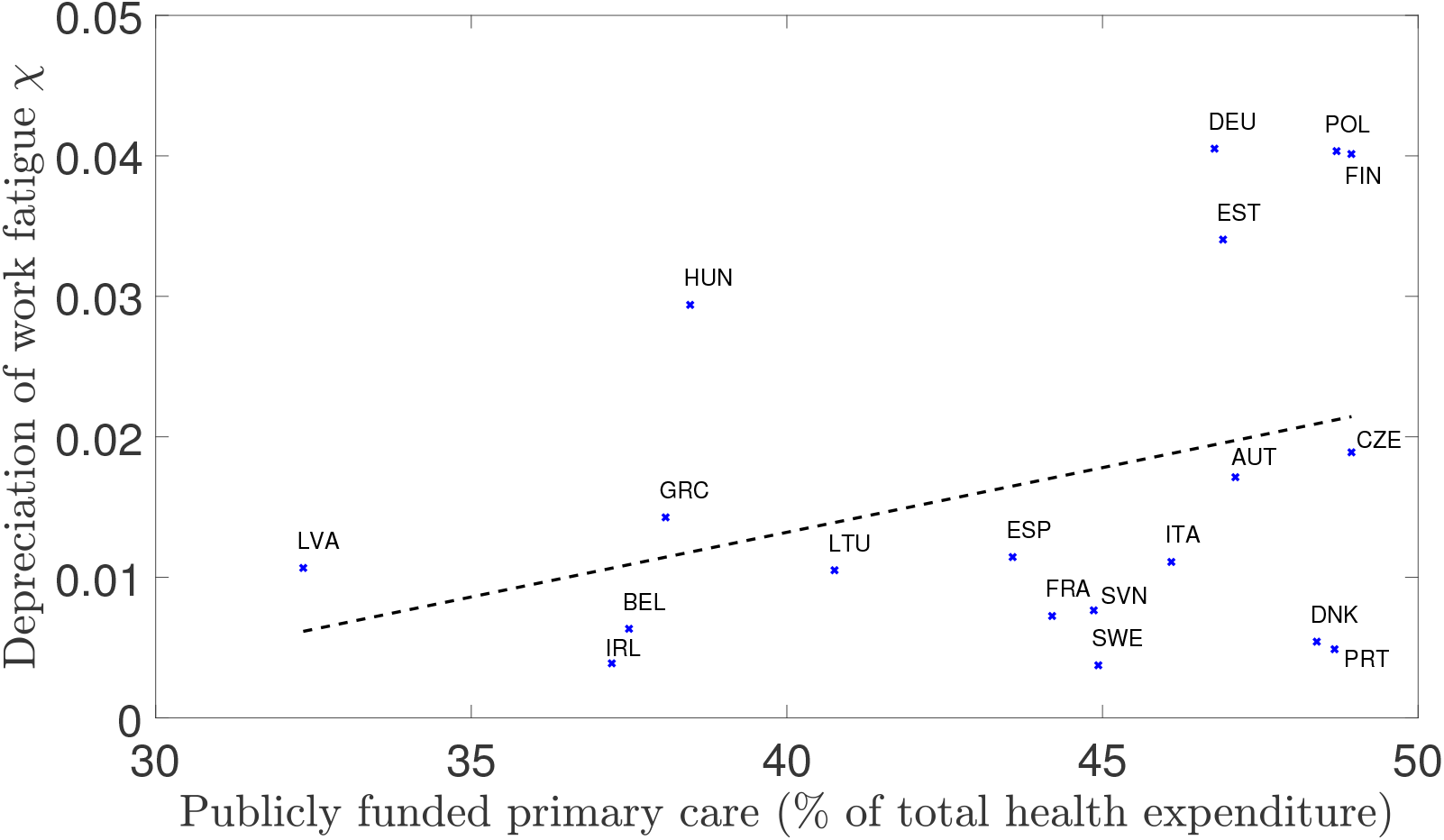
Calibrated values of *χ* can partly be explained by primary care public expenditure. The trend line shows the fitted values of a (simple) linear regression, the *p*-values are reported in parentheses. With slope/coefficient = 0.09 (*p* = 0.148), and intercept/constant = −0.02 (*p* = 0.390). The coefficient of determination is given by R^2^ = 0.12, and the sample size is *N* = 19.

Nevertheless, the positive relationship, and the size of the coefficient of determination, lend some support to the view that public provisions of healthcare services toward coping and recovery matter for work fatigue on aggregate. A more precise measure would consider only those provisions aimed at mental health or stress-related issues. However, we still take this apparent relationship as an indication that the distribution of depreciation rates from calibrating the model can partly be explained by empirical measures of public provisions of primary care services.

## 5 Discussion

In this section we discuss the differences between the baseline model detailed in this paper and the application of the previous section. In particular, we focus on the shortcomings associated with (i) comparing cross-section aggregate data using a time-series observation of a representative individual, and (ii) the issue with proxying fatigue accumulated from stressful work demands using sick leave rates from work stress.

Next, we extend the baseline model of Section 2 along two directions. First, we allow for wealth accumulation in a small-open economy setup. The possibility to invest into a saving technology to allow consumption smoothing while decreasing hours worked allows the individual to manage fatigue more efficiently. Second, we allow for human capital investments in addition to wealth accumulation. There, the individual can insure against increasing fatigue by becoming more productive, allowing them to reduce hours worked over time.

### 5.1 National-level cross-section vs individual time series

The baseline model describes the time-series behaviour of *N* identical individuals while Figure 5 shows a cross-section of country aggregates. This distinction matters as policy based on observations of individual workers may not translate easily into multiple country-level measures.

This apparent incompatibility between the two measures can of course be mitigated. Given the fact that the model is deterministic in nature, we could create heterogeneity in the baseline parameters of the model. For this, we could proceed in two ways. First, we could introduce heterogeneity in the preference parameters of the model, via the utility function in (6) using an appropriate distribution of parameters calibrated to data. Then, for a sufficiently large population size *N*, these population shares will correspond to individual probabilities of having a certain parameter set. This approach is most suited for capturing aggregate behaviour within country. Second, we could keep the preference parameters as they are, but instead vary only the policy and institutional parameters from the law of motion for fatigue in (7). Using this approach, we can replicate differences across nations by varying the underlying parameter set governing how hours worked affect the intertemporal utility costs of work, i.e. fatigue. Naturally, a combination of both can be used to match national-level cross-section data over time.

In the previous section, our aim was not to consider aggregate behaviour over time within country but merely to consider how the model could help us interpret the data at hand. Therefore our focus on varying the underlying parameters of equation (7) was the simplest move in that direction. Further research of interest could look at the evolution over time of the interplay between hours worked and fatigue measures within panel data if available. This could help us to test for the results and hypotheses of the baseline model.

### 5.2 Fatigue from work demands vs sick leave from work stress

Another difference between the baseline model explored in this paper and the application in the previous section is that the model makes use of work fatigue while the data shown used sick leave rates from work stress (inc. anxiety and depression from work-related activities). Evidently, the two types of measures are indeed different, and if nothing else, their supports are not identical, with one being defined on the positive real line while the other is constrained to the unit interval. However, we contend that the two measures are strongly positively correlated and that sick leave rates from work-stress are a good proxy for fatigue. It would specifically capture only the effect of fatigue above a certain threshold, as sick leave from work stress would only occur when the level of fatigue is high enough. This choice is motivated by the organisational psychology literature. Peterson *et al.* (2008) find that individuals with higher levels of work fatigue also tend to experience greater anxiety, while Hakanen and Schaufeli (2012) find that it correlates with depression. Additionally, this literature has also found strong effects of work fatigue on absenteeism, sick leave, and employee turnover (Lee and Ashforth, 1996; Wright and Cropanzano, 1998; Demerouti *et al.*, 2009). This body of evidence from psychology is not exhaustive and many studies have replicated those findings in various samples of the population, for multiple occupations and in several countries. We also return to the initial contribution, in economics rather than in psychology, by Banerjee *et al.* (2017), who, as mentioned earlier, find that declining mental health status is linked with increased sick leave rates.

One may ask whether the direction of the effect is that fatigue increases depression, anxiety, and sick leave or whether there is a degree of reverse causality. Another potential issue could be that a tertiary phenomenon is driving the pattern, or that those measures are simply cointegrated over time. However, even in this case, the use of sick leave rates from work-stress remains a valid proxy for fatigue from stressful work demands. Concerns over endogeneity or reverse causality would be a problem for regression analysis, however since the exercise here is purely to give a tentative explanation for a stylised fact, we do not yet have to be concerned with these issues. Another approach could be to measure the predictions of the model on hours worked as work intensity increases using panel data or time series of cross section survey data.

### 5.3 Wealth accumulation in a small open economy

The baseline model explored in this paper does not allow individuals to have access to an asset market and to shift consumption over time smoothly. It restricts individuals to consume only out of labour income, forcing consumption and labour supply to move one to one. This forces all dynamics of the model to be captured by work fatigue. In this section we extend this model to allow the individual consumer to save into an asset and accumulate wealth. Thus we now adopt a framework much closer to the standard Neoclassical model. We also abstract from ex-ante heterogeneity concerns. The introduction of ex-ante heterogeneity here would only force us to be a little more restrictive in the conditions required to obtain our results below. However, the aim of this section is only to verify that the essence of the model can survive theoretical extensions. Although not explicitly mentioned below, we will assume that the transversality condition holds. In this extension, the individual would face the following budget constraint, which replaces (4) with

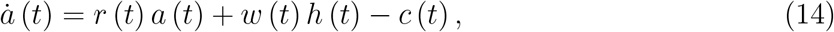

where *r* (*t*) ∈ ℝ_+_ is the rate of return on wealth *a* (*t*) ∈ ℝ, while the rest is as before. In this case, we would now be able to disentangle consumption and labour supply patterns, while studying the effects of wealth as a method of self insurance against increasing fatigue. The resulting optimal behaviour for the *N* identical individuals in the economy is now given by the following differential equations for consumption and labour supply^21^ (see app. A.4 for details),

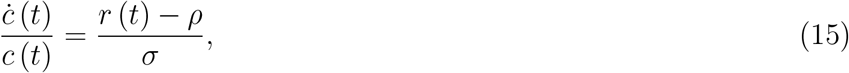

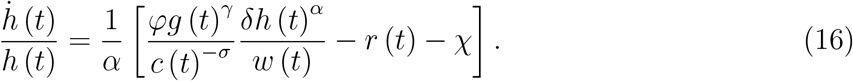

We see that the consumption rule is the standard Keynes-Ramsey rule for continuous time consumption-saving models, while the rule for labour supply is almost identical to the one in the model without wealth. The rate of return on wealth *r* (*t*) replaces *ρ* and *σ* no longer affects the curvature of the growth rate of labour supply. Given that wealth now exists in the economy, we can further extend the model by adding capital as an additional factor of production in the representative firm’s production technology, and opening up the economy to trade capital goods, while labour remains domestically bound. This small-open economy model means that factor rewards for capital will be given by *r*^*^ > 0, the international interest rate. This implies that, if the firm has a Cobb-Douglas production function, such as *Y* (*t*) = *AK* (*t*)^*θ*^ *L* (*t*)^1−*θ*^, with 0 < *θ* < 1 the share of capital in the production process, the capital to labour ratio *K* (*t*) */L* (*t*) will be fixed. This can be seen from the first-order condition of the firm for capital, giving us (upon rearranging)

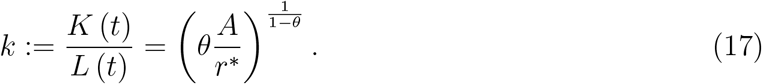

The main consequence of this result is that the wage rate *w* will also be constant, and given by the other first-order condition of the firm: *w* = (1 − *θ*) *Ak*^*θ*^, with the capital-labour ratio *k* being given by (17). In such a framework, the domestic level of capital is independent of domestic wealth holdings by households, due to the mobility of capital, and thus we abstract from linking aggregate capital to domestic wealth holdings.

In this extension of the baseline model, each individual would now be able to insure against fatigue over time by investing into wealth *a* (*t*), in order to fund future consumption. This pattern of behaviour would allow them to vary hours worked even more to reduce fatigue even faster. As there would no longer be a direct one-to-one drop in consumption, hours worked could drop more while the individual consumes out of their wealth in order to slow down the growth rate of, or even reduce, fatigue. In fact, along the balanced growth path [BGP], aggregate capital, labour, output, and consumption would all grow at the same rate *b* > 0.^22^ Capital and labour must grow at the same rate by the condition for the capital-labour ratio in (17). By linear homogeneity, output must also grow at the same rate, since technology is constant. And finally, by market clearing conditions, aggregate consumption must also grow at rate *b*. This implies that individual consumption and individual labour supply must also grow at that same rate, given that population is constant. It is then straightforward to show that, along this BGP, fatigue would have to be decreasing over time.^23^

#### Proposition 3

(Balanced growth recovery). *In the extended model with wealth accumulation, if all workers are identical and there is no heterogeneity, labour supply and fatigue will move in opposite directions for all workers, with*

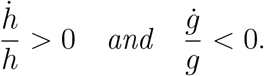

*Proof.* See app. A.5.

▪

Thus, along the BGP, we would observe a negative relationship between hours worked and fatigue, albeit one where hours worked actually increase over time and fatigue reduces. According to our phase diagram in Figure 1, this could happen if hours worked are low enough and fatigue is high enough. We could also make the argument that in order to observe declining hours worked over time along the BGP, the economy would need to contract, and consumption would need to drop over time. However, this assumption is incompatible with data. Of course, the introduction of positive growth rates of technology and population could shift these relationships and affect the results. At the individual level, becoming more productive could be a way to reconcile growing consumption schedules with decreasing paths of hours worked. This last point is explored in the second extension below.

However, before discussing the introduction of human capital it is worth noting that we may also simply depart from the BGP assumption. Instead, matching observations from data, we can note that aggregate consumption, measured by household spending, has been increasing in recent decades while aggregate hours worked have been falling (OECD, 2018b). If we assume constant growth rates for each process, then we obtain the following result.

#### Corollary 2.

*If consumption grows at* 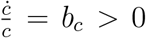, *and hours worked fall at* 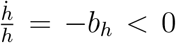, *fatigue will increase if and only if*,

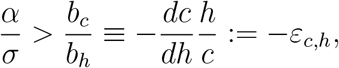

*where ε*_*c,h*_ *is the elasticity of consumption in labour supply.*

*Proof.* See app. A.6

▪

In words, as long as the elasticity of consumption in hours worked is small enough, we will observe a negative relationship between hours worked and work fatigue. Thus we see once more that the negative relationship between hours worked and work fatigue survives the introduction of wealth accumulation, even when the model does not follow the BGP.

In this situation, the capital stock will need to grow at the rate 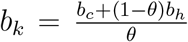 in order to make up for the declining path of labour supply while financing output growth. We note that in a small-open economy, this is not guaranteed, as the capital stock will depend partly on international inflows regulated by the international interest rate *r*^*^. Furthermore, with consumption increasing over time and labour supply falling, individual wealth would eventually run out. This would force individuals to either borrow in order to finance future consumption or to temporarily increase labour supply and reduce consumption to increase their wealth.

### 5.4 Human capital investment and self-insurance

As mentioned in the previous section, there exists another important determinant of earnings that has not been discussed which can affect individual productivity and allow individuals to further insure against the (utility) costs of high work intensity. Indeed, since the early work by Becker (1964) and Mincer (1974), we know that human capital investments are key to individual income streams. Furthermore, Mankiw *et al.* (1992), among others, have shown how important human capital accumulation is for growth at the country level.

In the context of the current paper however, we will consider human capital investments in order to enrich the income profile of individuals in the model and study the effects on labour supply choices. Thus, we consider the human capital of an individual at date *t* and call it *e* (*t*) ∈ ℝ_+_. This measure captures the efficiency of the individual in generating income when working, one may thus think of human capital in this context as in Lucas (1988). We can think of it as being the general skill level of the individual, accumulated either via schooling or on-the-job learning. We do ignore spillover effects of human capital accumulation. Such an effect could take place if the overall skill level of the economy were allowed to influence the individual skill level of the workers. We will also consider human capital as separate from the level of technology *A*.

The worker in this model will now face the following budget constraint, where human capital influences labour income such that the individual’s wealth will change with interest return on their investment plus earnings from efficiency units of labour 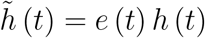 at the prevailing wage,

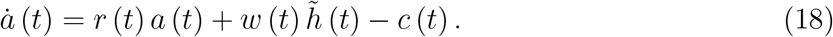

Furthermore, the individual can invest in human capital by choosing labour supply *h* (*t*) with return on investment *i* (*t*) > 0. This is captured by the function *v* (*h* (*t*)), which if it increases in *h* (*t*) would capture on-the-job learning and work experience, while if it decreases with *h* (*t*), we would consider it closer to schooling and education.^24^ Meanwhile the stock of existing human capital will depreciate (either by knowledge becoming irrelevant because of new technologies or because of imperfect recall of non-practiced skills) at the constant rate *η* > 0, giving us

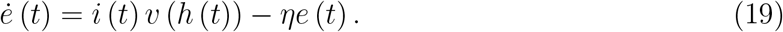

The rest of the consumer problem is as in the baseline model.

We also need to take into account that human capital will be used at the aggregate level to produce the final good. This is shown in the production function of the representative firm, now given by the following where the firm uses efficiency units of labour to produce *Y* (*t*)

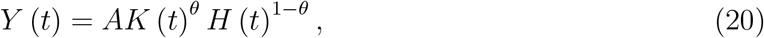

where aggregate efficiency units of labour *H* (*t*) is measured by 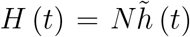.^25^ Thus once again, aggregate and individual variables will grow at the same rate. We again consider a small-open economy. The result is that, as before, factor rewards are now fixed and determined by the international interest rate *r*^*^, such that the wage is given by *w* = (1 − *θ*) *Aκ*^*θ*^, where *κ* := *K* (*t*) */H* (*t*) is the (constant) ratio of capital to efficiency units of labour. Once again, domestic wealth holdings and the domestic aggregate physical capital stock will be independent, as physical capital is internationally tradable.

Along the BGP, aggregate output, consumption, physical capital, and efficiency units of labour will all grow at the same rate *b* > 0. Given our definition of *H* (*t*), this implies that the growth rate of the economy is related to investment in human capital and the growth rate of labour supply by

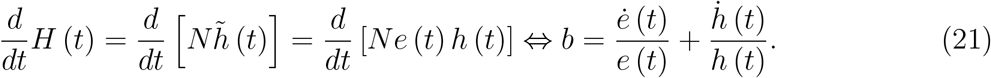

From the expression describing the evolution of human capital, we know that *e* (*t*) and *h* (*t*) can grow in opposite directions if *v′*(*h*) < 0. We can note here that, unlike in the model with only asset holdings, when human capital investment is introduced it is possible to have a balanced growth path, together with a declining path of hours worked. The growth rate of hours worked is then directly proportional to the growth rate of the economy, *b*.

#### Lemma 4.

*Along the BGP, and given the definition of aggregate efficiency units of labour (i.e. human capital* × *hours worked), the growth rate of hours worked will be constrained by the change in human capital stock and the growth rate of the economy, giving us the following identity for all workers*,

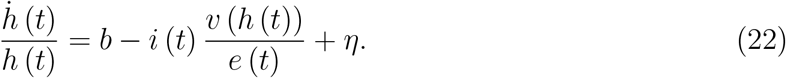

Thus, if the return on investment of human capital is high enough, we will observe falling hours worked. At the beginning of the planning horizon, when human capital is small, the individual has to work longer hours in order to build up their asset position. As wealth increases and human capital slowly builds up (still under the assumption that the returns are high enough), the individual is able to progressively work fewer hours. This is because they are able to enjoy the same labour income level for lower levels of labour supply due to the higher level of human capital. However, having started at a high level of hours worked, their work fatigue from exposure to stressful job demands will increase. This is because near the start of the planning horizon fatigue is also small, as the individual has not yet suffered from high work intensity. As hours worked progressively fall, as implied by Lemma 4 for high enough returns on investments in *e* (*t*), the rate at which fatigue grows will slow down but remain positive until hours worked become low enough to enable the individual to recover. Whether the individual ever arrives at this point depends on how productive investments in human capital are and how quickly knowledge becomes obsolete. It is also possible for the individual to experience cycles of working long hours, accumulating human capital, then reducing their labour supply until their human capital has fallen enough to force them to increase their labour supply once again. Along these cycles, fatigue would wave counter-cyclically, following the profile of hours worked with a delay.

## 6 Conclusion

In this paper, we have explored the role of work intensity, as measured by stressful work demands, on hours worked. High work intensity causes intertemporal utility costs for the individual via their choice of hours worked, which we call “fatigue”. The model developed here reconciles some observations made on the pattern of hours worked over time and aggregate measures of fatigue caused by work stress. This gives rise to a behaviour whereby individuals work long hours when the utility costs are still low, before decreasing labour supply as fatigue settles in as a result of their past labour supply decisions.

We take the view that this mechanism is an important one in the determination of labour supply choices for individuals. We apply this framework to a stylised observation from data that aggregate hours worked and sick leave rates from work stress are negatively correlated in a cross-section of European countries. The institutional background on healthcare services and the share of services in each country is taken as the key variation across economies, and the calibration exercise offers evidence to support that view.

We discuss the potential limitations of this application as well as two extensions of the baseline model. The first extension accounts for wealth accumulation and shows that even when we can disentangle consumption and hours profiles, the negative relationship between hours worked and fatigue holds along the balanced growth path. For consumption and hours profiles closer to real-world observations, the main result survives and hours worked can be observed to fall while fatigue increases over time. The second extension introduces the possibility to invest into human capital in addition to wealth. In this version of the model, individuals can now insure against increasing fatigue via asset holdings or higher productivity to finance future consumption. The main result of the baseline model, namely that hours worked and fatigue are negatively correlated, remains valid along the balanced growth path as long as returns on investment in human capital are high enough.

## Data Availability

The data used in the final section of the paper comes from Eurostat and th OECD databases.

## A Appendix

### A.1 Optimal Behaviour

We employ the Hamiltonian approach to solve the optimisation problem of the consumer. Using (2), (4), and (1), we can now use *h*_*i*_ (*t*) as our control variable. Note that in the steps that follow, we write the wage rate as being time dependent. However, in the main text, the wage rate is constant by construction. This allows us to have more general expressions here while the main text presents a special case. We begin by writing the Hamiltonian

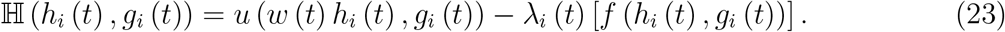

Optimality conditions for this problem are then given by the first-order condition

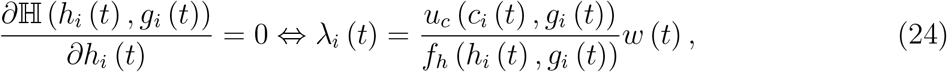

and the following differential equation for the co-state variable,

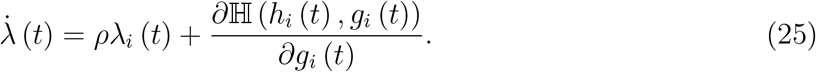

We can solve this system by taking the derivative of (24) w.r.t. time and then replacing for *λ* (*t*) and 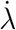 (*t*) in (25), and rearranging. The solution gives us the dynamic rule for hours worked,

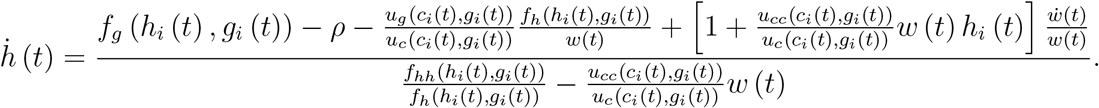

### A.2 Dynamics of Motion

We can derive the laws of motion for *g*_*i*_ (*t*) and *h*_*i*_ (*t*) using (6) and (7). We can rewrite the system of dynamic equations above as follows, and holding the wage rate constant, as in the main text,

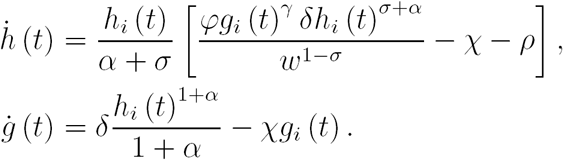

In order to understand the dynamics of the system, we look at the zero-motion lines defined as

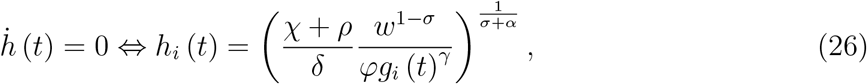

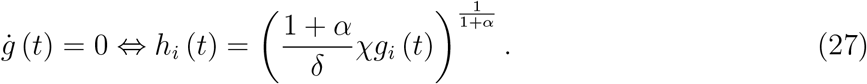

We also need to check the asymptotic behaviour of both zero-motion lines, thus looking at the system above we have that

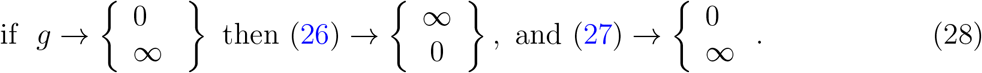

Furthermore, we need to understand how each function behaves between those limit values, by checking their first and second-order derivatives, omitting time arguments for simplicity,

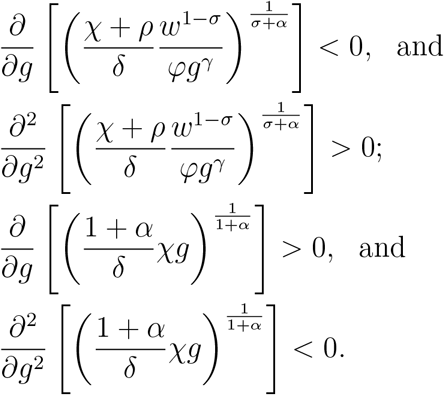

Therefore we have that the zero-motion line for *ġ* (*t*) will be increasing concave in *g*_*i*_ (*t*), while the one for 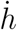 (*t*) will be decreasing convex in *g*_*i*_ (*t*). Finally, we now need to understand how each variable moves outside of the zero-motion lines, to determine the path to equilibrium, so that given

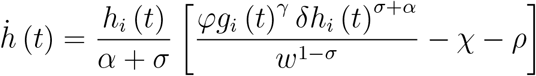

we have the following condition that labour supply *h*_*i*_ (*t*) grows when it lies above its zero-motion line

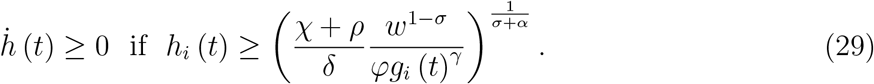

Meanwhile, given

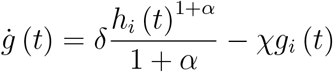

we have the following condition that fatigue *g*_*i*_ (*t*) grows whenever labour supply is above the zero-motion line for fatigue,

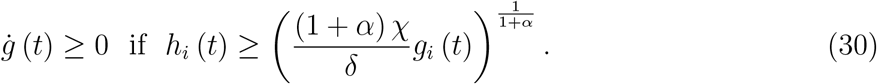

### A.3 Transversality Condition

Using our first order condition from the Hamiltonian in Appendix A.1, we can check that the TVC in (13) holds, by first plugging for *λ* using the first-order condition in equation (24),

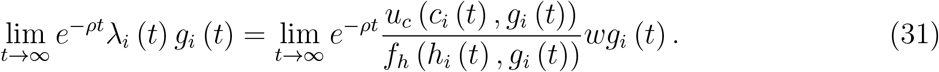

Given the domain of *h*_*i*_ (*t*), we know that *g*_*i*_ (*t*) will be at its highest when 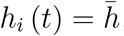, ∀*t*, which we denote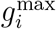. We can write

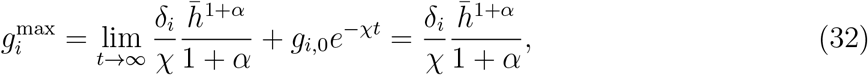

which we can use to write the following inequality for any other path of *h*_*i*_ (*t*),

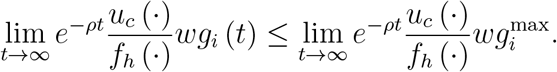

The right-hand side of the expression above then provides an upper bound on the TVC. Now using (6)-(7) and (32) above, the expression on the right-hand side above reads

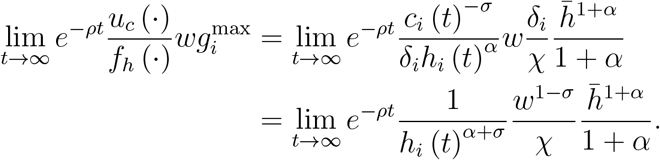

Given that the solution to the Hamiltonian in Appendix A.1 yields a strictly interior solution for *h*_*i*_ (*t*), and as long as wages are bounded, we have the following limit as *t* goes to infinity,

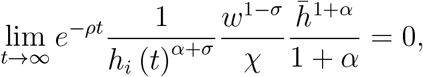

which bounds our TVC since,

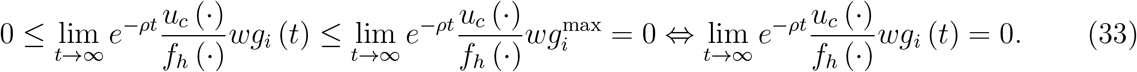

The last equality holding by definition, since the terms in the TVC are all strictly non-negative. Thus, the TVC holds in equilibrium, so long as wages do not go to infinity, which is trivially satisfied in a small-open economy setup, as factor rewards are constant.

### A.4 Optimal Rules with Wealth Accumulation

We first write the Bellman equation of the model with wealth, omitting time arguments,

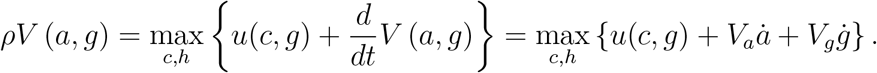

The first-order conditions of the model are then given by

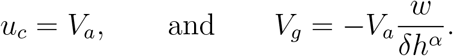

We can write the evolution of the co-state variables by taking the derivative of the maximised Bellman equation above, w.r.t. each state variable. This step gives us

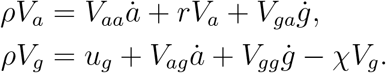

We can also express the evolution of the co-state variables by taking the derivatives of each co-state w.r.t. time,

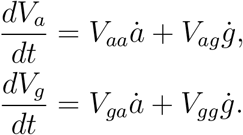

Combining our two results, we obtain the following expressions

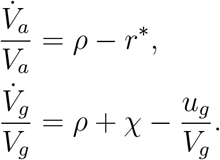

Next, we can insert the FOCs and rewrite the resulting expressions to obtain the optimal dynamic rules,

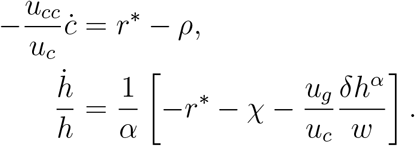

Plugging the corresponding expressions for the derivatives of the utility function then yields the expressions seen in the main text.

### A.5 Proof of Proposition 3

We note that along the BGP, aggregate variables grow at the same rate. Here, since aggregate and individual level variables grow at the same rate, given that all individuals are the same and there is no heterogeneity, we can use our dynamic rules to obtain our condition for growth in fatigue.

Setting, 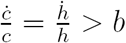, where *b* > 0 is the constant growth rate along the BGP, we obtain

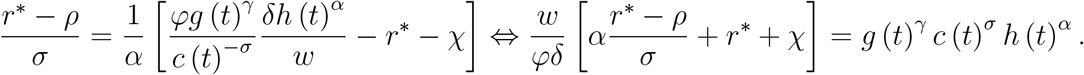

Thus we have the condition that the expression above needs to hold along the BGP. For this to be true, we need the time derivative of the right-hand side to be equal to 0. Thus, taking the derivative of this condition w.r.t. time and noting that we have an economy of identical individuals with no heterogeneity,

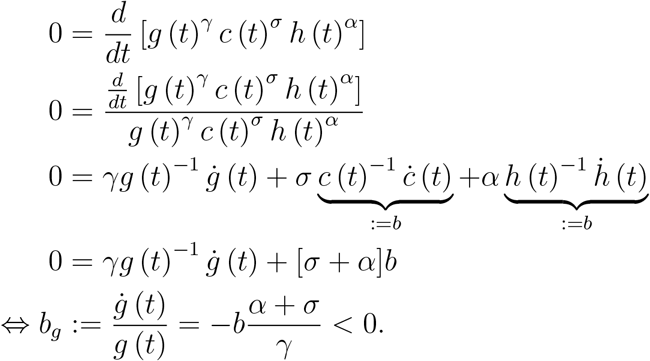

### A.6 Proof of Corollary 2

Consider a small-open economy where workers accumulate work fatigue as a consequence of their labour supply decisions, and they have access to a saving technology. Assume consumption and labour supply each evolve over time according to 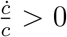 and 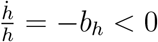, where *b*_*c*_and *b*_*h*_ are constant rates.

Then, we can write

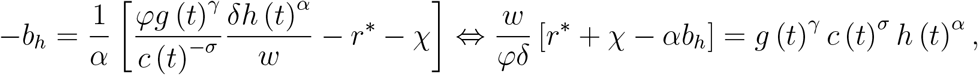

which has to be constant. Thus, just as before, the derivative of this expression w.r.t. time must be equal to 0, the key difference being that this time the growth rates will not be identical for consumption and labour supply. We obtain the following condition

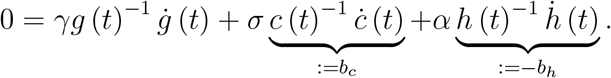

Rearranging gives us,

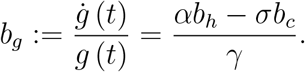

And to obtain the result mentioned in Corollary 2, we can simply see the condition required to have *b*_*g*_ > 0. This condition reads,

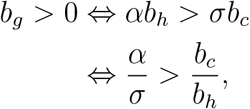

where we finally note that the ratio of the growth rates can be re-written as

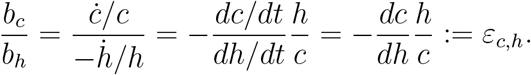

### A.7 The Model with Wealth and Human Capital

We first write the Bellman equation of the model with wealth, omitting time arguments,

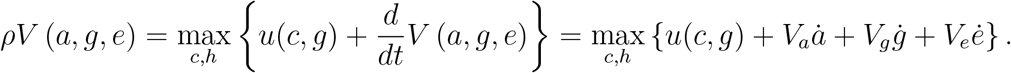

The first-order conditions of the model are then given by

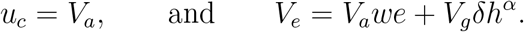

We can write the evolution of the co-state variables by taking the derivative of the maximised Bellman equation above, w.r.t. each state variable. This step gives us

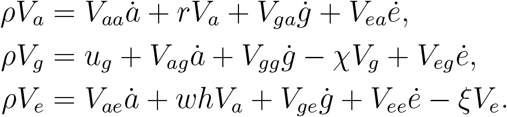

We can also express the evolution of the co-state variables by taking the derivatives of each co-state w.r.t. time

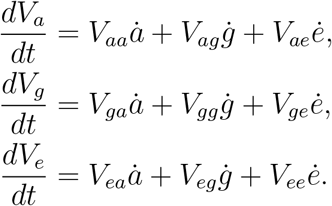

Combining our two results, we obtain

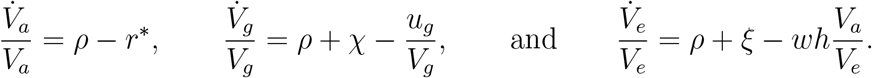

Combining these expressions with the FOCs from above, we obtain the system of equations that characterise the individual problem in equilibrium.

## Footnotes

1 Kessler *et al.* (2008) provide a survey of the literature on the epidemiology of mental health issues for the World Health Organization.

2 The measure used to capture sick leave from work stress includes work-related stress, anxiety, and depression resulting in sick leave as a share of total sick leave in that year (see Labour Force Survey, 2017).

3 Early work by Yaniv (1995) looks at the effect of absenteeism and stress on overtime decision. Empirically, Golden and Wiens-Tuers (2006) look at the effect of overtime work on measures of well-being, while Hamermesh and Lee (2007) look at the importance and prevalence of time stress.

4 The effects of savings and human capital investments are explored in Section 5.

5 In the organisational psychology literature, the leading frameworks to conceptualise recovery are the EffortRewards Imbalance and Effort-Recovery models (Meijman and Mulder, 1998; Siegrist *et al.*, 2004). The takeaway is that individuals must recover proportionately from fatigue while being compensated accordingly in order to avoid severe episodes of discontent and lower levels of well-being.

6 While psychologists do not make the point explicitly, it is well documented that work fatigue and burnout, while present across several occupation types and sectors, are most prevalent in the service sector. Thus, we merely bring an economist’s view to psychological observations.

7 We can note here that both GHQ scores and other measures of fatigue, e.g. ERI scores, tend to increase with age (see O’Connor and Parslow 2010). Thus, there may be an additional channel over time that we are currently ignoring that may be driving some of the downward trend in hours worked at older ages.

8 A small-open economy setup is explored in later sections, however we abstract from the effects of trade between countries.

9 This assumption of a constant productivity level *A* may seem restrictive at first. However, we note here that its implication, namely to have constant wages, is similar to what would happen in a small open economy with capital inputs. In that case, the international rate of return *r*^*^ would be internationally fixed, which would also fix the wage rate by forcing the capital-labour ratio to be constant, whenever we have linear homogeneity of degree one in production (e.g. CES type functions). In such a framework, the level of domestic capital would be independent of wealth held by domestic households. This type of economy is discussed in the extensions of the model.

10 This assumption is relaxed later in Section 5, and its implications are discussed.

11 The wage is constant, in accordance with the result of the previous section and the optimal behaviour of the representative firm since *A* is also constant and *L* (*t*) is the only factor of production.

12 We can see this through Lemma 3.(i.d), where we can take the derivative of (10) w.r.t. *δ*_*i*_ to obtain the condition 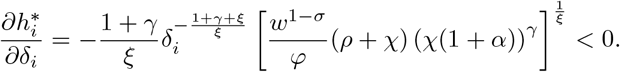

13 See Kamihigashi (2006) and Hestenes (1966) for a more complete discussion on the necessity of TVCs in economics and in optimal control problems in general, respectively.

14 Similar arguments can be made for the remaining pairs of aggregate and individual variables.

15 To move from a continuous time preference rate to an annual rate of discounting, we simply see that the following must hold for any bundle *x*_0_: *x*_0_(1 + *ρ*_*y*_) = *x*_0_(1 + *ρ*_*m*_)^12^, where *ρ*_*y*_ and *ρ*_*m*_ are yearly and monthly discount factors. Meanwhile, if the value of this bundle evolves over time according to 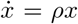, then we must also have *x*_0_*e*^*ρ*^ = *x*_0_(1 + *ρ*_*m*_)^*s*^, where *s* is the number of periods in one unit of time, as captured by the model’s time preference rate *ρ*. Here if one unit of time is one year, then moving from month to year requires 12 periods, and thus *s* = 12. Simple algebra then allows us to obtain the following equivalence: 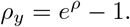

16 This value may appear high, however it is assuming the level of work fatigue of the individual is at its lowest possible point on the ERI scale, which is unlikely to occur in the real-world.

17 We note here that the implied income elasticity of labour is 0.15 using the parameter set of the numerical solution. This value is in line with empirical estimates (for details see Keane *et al.*, 2011). We do not contend that this implied elasticity is in any way relevant for empirical work, but merely make note that the parameter set, both calibrated and exogenous, leads to a value of *ε*_*h*\*_,*w* that appears reasonable. This adds to our confidence in the results obtained in this paper. Of course, we understand the caveat that this result is conditional on observing a steady-state, which has its drawbacks and does not necessarily correspond to elasticities in transitional dynamics.

18 We abstract from giving explicit interpretations to these values and focus instead only on their relative change.

19 We are aware of the limitations of fitting a linear trend between levels of long-run work fatigue and observed sick leave rates, but the aim of this numerical exercise is simply to gain some insights into what the model can tell us about the data, using the tools at our disposal.

20 The graph very clearly shows that there are five outliers, lying north of the other countries, with *χ* values larger than 0.03. Separating the sample into two sub-samples yields a (nearly) flat and insignificant slope for countries with *χ* < 0.02 while it does yield a positive significant slope for countries with *χ* > 0.03. However, the second sample comprises only five countries and therefore lacks credibility regarding the significance of the relationship. For this reason, we abstract from including it in the main text.

21 Aggregate consumption *C* (*t*) = *Nc* (*t*) grows at the same rate as the sum of changes of each individual consumption *Ċ* (*t*) = *N ċ* (*t*), other aggregate growth rates are defined analogously.

22 See Turnovsky (2009) for a more extensive discussion of balanced growth paths in small-open economies.

23 We can show this by setting the condition that 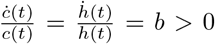, using the dynamic rules above, and then rearranging to show that fatigue needs to fall at rate 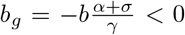, in order for this condition to be true. The steps do rely on *r*^*^ and *w* being constant, as implied by the small-open economy setup.

24 In the context of this model, education is understood as a continuous choice by the individual alongside hours worked. One possible explanation is that individuals participate in summer schools, intensive programmes, or professional certifications in order to gain new skills. Doing so happens at the cost of hours spent working. We thus differ from the usual definition of education as occurring before the working life of the individual.

25 Once again we exploit the fact that in a deterministic model with no heterogeneity in parameter and constant population, the aggregate variables are simply the sum of individual variables over all *N* workers.

## References

Andersen, S., G. Harrison, M. Lau, and E. Rutström (2008): “Eliciting Risk and Time Preferences,” Econometrica, 76(3), 583–618.

Avgoustaki, A., and J. Frankort (2019): “Implications of work effort and discretion for employee well-being and career-related outcomes: an integrative assessment,” Industrial and Labor Relations Review, 72(3), 636–661.

Banerjee, S., P. Chatterji, and K. Lahiri (2017): “Effects of Psychiatric Disorders on Labor Market Outcomes: A Latent Variable Approach Using Multiple Clinical Indicators,” Health Economics, 26(2), 184–205.

Becker, G. (1964): Human Capital: A Theoretical and Empirical Analysis with Special Reference to Education. Chicago, IL: University of Chicago Press.

Becker, G. (1965): “A Theory of the Allocation of Time,” Economic Journal, 75, 493–517.

Blinder, A., and Y. Weiss (1976): “Human Capital and Labor Supply: A Synthesis,” Journal of Political Economy, 84, 449–472.

Bundesanstalt für Arbeitsschutz und Arbeitsmedizin (2016): Sicherheit und Gesundheit bei der Arbeit 2015: Unfallverhütungsbericht Arbeit. Dortmund: Bundesanstalt für Arbeitsschutz und Arbeitsmedizin.

Cygan-Rehm, K., and C. Wunder (2018): “Do working hours affect health? Evidence from statutory workweek regulations in Germany,” Labour Economics, 53(August), 162–171.

Demerouti, E., A. Bakker, F. Nachreiner, and W. Schaufeli (2001): “The job demands-resources model of burnout,” Journal of Applied Psychology, 86, 499–512.

Demerouti, E., P. Le Blanc, A. Bakker, W. Schaufeli, and J. Hox (2009): “Present but sick: a three-wave study on job demands, presenteeism and burnout,” Career Development International, 14(1), 50–68.

Erosa, A., L. Fuster, and G. Kambourov (2016): “Towards a Micro-Founded Theory of Aggregate Labor Supply,” Review of Economic Studies, 83(3), 1001–1039.

Eurofound (2010): “Absence from work,” European Foundation for the Improvement of Living and Working Conditions.

French, E. (2005): “The Effects of Health, Wealth, and Wages on Labour Supply and Retirement Behaviour,” Review of Economic Studies, 72(2), 395–427.

Golden, L., and B. Wiens-Tuers (2006): “To your happiness? Extra hours of labor supply and worker well-being,” Journal of Socio-Economics, 35, 382–397.

Gourinchas, P., and J. Parker (2002): “Consumption Over the Life Cycle,” Econometrica, 70(1), 47–89.

Grossman, M. (1972): “On the Concept of Health Capital and the Demand for Health,” Journal of Political Economy, 80(2), 223–255.

Hakanen, J., and W. Schaufeli (2012): “Do burnout and work engagement predict depressive symptoms and life satisfaction? A three-wave seven-year prospective study,” Journal of Affective Disorders, 141(2), 415–424.

Hamermesh, D., and J. Lee (2007): “Stressed out on Four Continents: Time Crunch or Yuppie Kvetch,” Review of Economics and Statistics, 89(2), 374–383.

Heckman, J. (1974): “Life Cycle Consumption and Labor Supply: An Explanation of the relationship between Income and Consumption Over the Life Cycle,” American Economic Review, 64(1), 188–194.

Heckman, J. (1976): “Estimates of a Human Capital Production Function Embedded in a Life-Cycle Model of Labor Supply,” in Household Production and Consumption, ed. by N. Terleckyj, pp. 225–264. NBER.

Heckman, J., and T. MaCurdy (1980): “A Life Cycle Model of Female Labour Supply,” Review of Economic Studies, 47(1), 47–74.

Heckman, J., and R. Willis (1977): “A Beta-logistic Model for the Analysis of Sequential Labor Force Participation by Married Women,” Journal of Political Economy, 85, 27–58.

Hestenes, M. (1966): Calculus of Variations and Optimal Control Theory. New York: John Wiley and Sons.

Hinz, A., M. Zenger, E. Braehler, S. Spitzer, K. Scheuch, and R. Seibt (2016): “Effort-Reward Imbalance and Mental Health Problems in 1074 German Teachers, Compared with Those in the General Population,” Stress and Health, 32(3), 224–230.

Kamihigashi, T. (2006): “Transversality Conditions and Dynamic Economic Behavior,” The New Palgrave Dictionary of Economics, 2nd Edition.

Kessler, R., and T. üstün(eds.) (2008): The WHO world mental health surveys. Global perspectives of mental health surveys. New York: Cambridge University Press.

Lang, M., C. McManus, and G. Schaur (2019): “The effects of import competition on health in the local economy,” Health Economics, 28(1), 44–56.

Layard, R. (2016): “The economics of mental health,” IZA World of Labor.

Lee, R., and B. Ashforth (1996): “A Meta-Analytic Examination of the Correlates of the Three Dimensions of Job Burnout,” Journal of Applied Psychology, 81(2), 123–133.

Lucas, R. (1988): “On the mechanics of economic development,” Journal of Monetary Economics, 22, 3–42.

Mankiw, N., D. Romer, and D. Weil (1992): “A Contribution to the Empirics of Economic Growth,” Quarterly Journal of Economics, 107(2), 407–437.

Meijman, T., and G. Mulder (1998): “Psychological aspects of workload,” in Handbook of work and organizational psychology, ed. by P. Drenth, and H. Thierry, vol. 2, pp. 5–33. Hove: Psychology Press.

Mincer, J. (1974): Schooling, Experience and Earnings. New York, NY: National Bureau of Economic Research.

O’Connor, D., and R. Parslow (2010): “Mental health scales and psychiatric diagnoses: Responses to GHQ-12, K-10 and CIDI across the lifespan,” Journal of Affective Disorders, 121(3), 263–267.

OECD (2012): Sick on the Job? Myths and Realities about Mental Health and Work, Mental Health and Work. OECD Publishing, Paris.

OECD (2014): Making Mental Health Count: The Social and Economic Costs of Neglecting Mental Health Care, OECD Health Policy Studies. OECD Publishing, Paris.

OECD (2018a): “Earnings: Average annual wages,” OECD Employment and Labour Market Statistics (database).

OECD (2018b): “Hours Worked: Average annual hours actually worked,” OECD Employment and Labour Market Statistics (database).

OECD (2019a): “Health spending,” doi: 10.1787/8643de7e-en.

OECD (2019b): “Value added by activity,” doi: 10.1787/a8b2bd2b-en.

Office of National Statistics (2017): “Sickness absence in the UK labour market: 2016,” https://www.ons.gov.uk/employmentandlabourmarket/peopleinwork/labourproductivity/articles/sicknessabsenceinthelabourmarket/2016.

Peterson, U., E. Demerouti, G. Bergström, M. Samuelson, M. Asberg, and A. Nygren (2008): “Burnout and physical and metal health among Swedish healthcare workers,” Journal of Advanced Nursing, 62, 84–95.

Roelen, C., M. van Hoffen, J. Groothoff, J. de Bruin, W. Schaufeli, and W. Van Rhenen (2015): “Can the Maslach Burnout Inventory and Utrecht Work Engagement Scale be used to screen for risk of longterm sickness absence?,” International Archives of Occupational and Environmental Health, 88, 467–475.

Schaufeli, W., M. Leiter, and C. Maslach (2008): “Burnout: 35 years of research and practice,” Career Development International, 14(3), 204–220.

Siegrist, J., S. Starke, T. Chandola, I. Godin, M. Marmot, I. Niedhammer, and R. Peter (2004): “The measurement of effort-reward imbalance at work: European comparisons,” Social Science and Medicine, 58, 1483–1499.

Statistical Office of the European Communities (2017): Eurostat, EU Health and Safety at Work : Persons reporting a work-related health problem by sex, age and type of problem. Luxembourg: Eurostat.

Turnovsky, S. J. (2009): Capital Accumulation and Economic Growth in a Small Open Economy. Cambridge, UK: Cambridge University Press.

Weiss, Y., and R. Gronau (1981): “Expected Interruptions in Labour Force Participation and Sex-Related Differences in Earnings Growth,” Review of Economic Studies, 48, 607–619.

Wright, T., and R. Cropanzano (1998): “Emotional Exhaustion as a Predictor of Job Performance and Voluntary Turnover,” Journal of Applied Psychology, 83(3), 486–493.

Yaniv, G. (1995): “Burnout, absenteeism, and the overtime decision,” Journal of Economic Psychology, 16, 297–309.

